# Third and fourth vaccine doses broaden and prolong immunity to SARS-CoV-2 in immunocompromised adult patients

**DOI:** 10.1101/2023.03.01.23286513

**Authors:** Michelle W Cheung, Roya M Dayam, Janna R Shapiro, Jaclyn C Law, Gary YC Chao, Daniel Pereira, Rogier L Goetgebuer, David Croitoru, Joanne M Stempak, Lily Acheampong, Saima Rizwan, Jenny D Lee, Liz Jacob, Darshini Ganatra, Ryan Law, Victoria E. Rodriguez-Castellanos, Madeline Kern-Smith, Melanie Delgado-Brand, Genevieve Mailhot, Nigil Haroon, Robert D. Inman, Vincent Piguet, Vinod Chandran, Mark S Silverberg, Tania H Watts, Anne-Claude Gingras

**Affiliations:** Department of Immunology, University of Toronto, Toronto, Ontario, Canada; Lunenfeld-Tanenbaum Research Institute at Mount Sinai Hospital, Sinai Health, Toronto, Ontario, Canada; Psoriatic Arthritis Program, Schroeder Arthritis Institute, Krembil Research Institute, University Health Network, Toronto, Ontario, Canada; Zane Cohen Centre for Digestive Diseases, Division of Gastroenterology, Mount, Sinai Hospital, Sinai Health, Toronto, Ontario, Canada; Division of Rheumatology, Department of Medicine, University of Toronto, Toronto, Ontario, Canada; Division of Dermatology, Department of Medicine, University of Toronto, Toronto, Ontario, Canada; Division of Dermatology, Department of Medicine, Women’s College Hospital, Toronto, Ontario, Canada; Laboratory Medicine and Pathobiology, University of Toronto, Toronto, Ontario, Canada; Division of Gastroenterology, Department of Medicine, University of Toronto, Toronto, Ontario, Canada; Department of Molecular Genetics, University of Toronto, Toronto, Ontario, Canada

## Abstract

**Background:** Previous studies have reported impaired humoral responses after SARS-CoV-2 mRNA vaccination in immunocompromised patients with immune-mediated inflammatory diseases (IMID), particularly those treated with anti-tumor necrosis factor (TNF) biologics. We previously reported that IMID patients exhibited greater waning of antibody and T cell responses compared to healthy controls after dose 2. Fewer data are available on the effects of third and fourth doses.

**Methods:** This observational cohort study collected plasma and peripheral blood mononuclear cells from healthy controls and untreated or treated IMID patients, pre-vaccination and after one to four doses of SARS-CoV-2 mRNA vaccine (BNT162b2 or mRNA-1273). SARS-CoV-2- specific antibody levels, neutralization, and T cell cytokine responses were measured against Wildtype (WT) and BA.1 and BA.5 variants of concern (VOCs).

**Results:** Third vaccine doses substantially restored and prolonged antibody and T cell responses in IMID patients and broadened responses against VOCs. Fourth dose effects were subtle but also prolonged antibody responses. However, IMID patients treated with anti-TNF, especially inflammatory bowel disease (IBD) patients, exhibited lower antibody responses even after the fourth dose. Although T cell IFNγ responses were maximal after one dose, IL-2 and IL-4 production increased with successive doses, and early production of these cytokines was predictive of neutralization responses at 3-4 months post-vaccination.

**Conclusion:** Our study demonstrates that third and fourth doses of the SARS-CoV-2 vaccine sustain and broaden immune responses to SARS-CoV-2, supporting the recommendation for three- and four-dose vaccination regimens in IMID patients.

**Funding:** COVID-19 Immunity Task Force and Speck family donation

**Conflict-of-Interest Statements:** Anne-Claude Gingras has received research funds from a research contract with Providence Therapeutics Holdings, Inc., for other projects, participated in the COVID-19 Immunity Task Force (CITF) Immune Science and Testing working party, chaired the CIHR Institute of Genetics Advisory Board, and chairs the SAB of the National Research Council of Canada Human Health Therapeutics Board. Vinod Chandran has received research grants from AbbVie, Amgen, and Eli Lilly and has received honoraria for advisory board member roles from AbbVie, Amgen, BMS, Eli Lilly, Janssen, Novartis, Pfizer, and UCB. His spouse is an employee of AstraZeneca. Vincent Piguet has no personal financial ties with any pharmaceutical company. He has received honoraria for speaker and/or advisory board member roles from AbbVie, Celgene, Janssen, Kyowa Kirin Co. Ltd, LEO Pharma, Novartis, Pfizer, Sanofi, UCB, and Union Therapeutics. In his role as Department Division Director of Dermatology at the University of Toronto, Dr. Piguet has received departmental support in the form of unrestricted educational grants from AbbVie, Bausch Health, Celgene, Janssen, LEO Pharma, Lilly, L’Oréal, NAOS, Novartis, Pfizer, Sandoz and Sanofi in the past 36 months. Vincent Piguet has received research grants from Sanofi, Abbvie and Novartis. Mark Silverberg has received research support, consulting fees and speaker honoraria from AbbVie, Janssen, Takeda, Pfizer, Gilead, and Amgen. All other authors have no conflicts to declare.

**Graphical Abstract:** 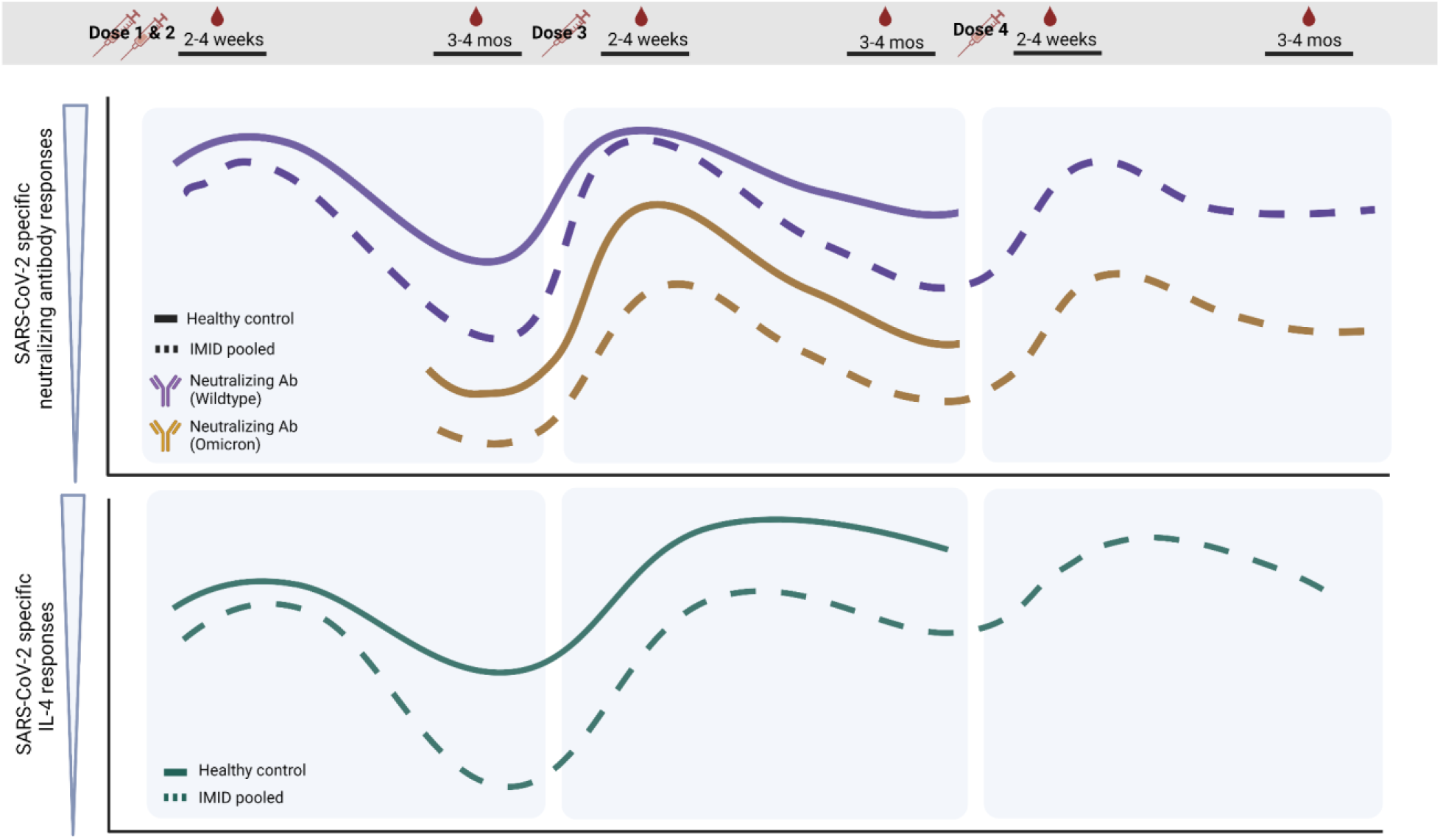

## Introduction

With the frequent emergence of new severe acute respiratory syndrome coronavirus 2 (SARS-CoV-2) variants, the coronavirus disease 2019 (COVID-19) pandemic remains a pressing global health concern (1). Patients with immune-mediated inflammatory diseases (IMID) have dysregulated immune systems, are often treated with immune-modifying medications, and are predisposed to higher risks of SARS-CoV-2 infection and severe outcomes following infection, including critical care admission and death (2, 3).

The initial clinical trials of the SARS-CoV-2 mRNA vaccines (BNT162b2 Pfizer/BioNTech and mRNA-1273 Moderna) excluded immunocompromised patients (4, 5), raising concerns about how to optimally protect these vulnerable patients. There is now substantial evidence to suggest that certain immunomodulatory therapies, such as B-cell depleting therapies, glucocorticoids, tumor necrosis factor (TNF) inhibitors, mycophenolate mofetil, JAK inhibitors, and methotrexate (MTX), can attenuate humoral and cellular responses to the primary series (2 doses) of SARS-CoV-2 mRNA vaccination (6–13). Data are lacking regarding the immunogenicity of the SARS-CoV-2 mRNA vaccines in immunocompromised IMID patients after repeated vaccination doses. Moreover, many studies primarily focused on humoral responses post-vaccination. However, adaptive immunity to SARS-CoV-2 depends not only on antibody (Ab) and neutralization responses, but on concomitant T-cell mediated responses (14–19). To our knowledge, there is a paucity of studies that longitudinally examine both humoral and cellular immunogenicity of SARS-CoV-2 mRNA vaccines in IMID patients throughout a primary series of vaccination and booster (third and fourth) doses. These data are necessary to inform the optimal vaccination strategy for this vulnerable population.

To further the knowledge of the impact of immunomodulatory drugs on the quality and magnitude of SARS-CoV-2 vaccine-induced immunity, we recently investigated the <u>Im</u>mune res<u>P</u>onse <u>A</u>fter <u>C</u>OVID-19 vaccination during maintenance <u>T</u>herapy in immune-mediated inflammatory diseases (IMPACT) by prospectively following a cohort of vaccinated IMID patients with inflammatory bowel disease, rheumatic or skin disease on immunosuppressive therapies compared to healthy controls (20, 21). We demonstrated that IMID patients exhibit greater waning of Ab and T cell responses to SARS-CoV-2 by 3-4 months post dose 2 compared to healthy controls (20). Notably, anti-TNF treated patients showed the greatest reductions in Ab responses in our cohort, had reduced efficacy of neutralization of variants of concern (VOCs), and could not neutralize Omicron BA.1 (20). TNF is critical for proper lymphoid-organ architecture and organization of germinal centers (GCs) (22, 23). The GCs, in turn, are needed to generate high-affinity antibodies, long-lived plasma cells and memory B cells. In the context of SARS-CoV-2, we and others have reported impaired Ab and neutralization responses to a primary series of vaccination in anti-TNF-treated IMID patients (6, 7, 10, 20, 24). This indicated that IMID patients treated with TNF inhibitors should be closely monitored over time for loss of humoral immunity to SARS-CoV-2.

There is limited data on whether third and fourth doses of SARS-CoV-2 vaccines can correct the deficits in immune responses of anti-TNF treated patients after 2 doses of vaccine, and whether successive boosters will augment the magnitude and durability of immunity to SARS-CoV-2 in healthy and IMID populations. To this end, here we report an extension of the IMPACT study to investigate the immunogenicity of third and fourth vaccine doses. We measured spike- (S) and RBD-specific Ab levels, neutralization and T cell cytokine responses at 2-4 weeks and 3-4 months after each booster dose of vaccine. The primary aim was to evaluate the effect of booster doses on the magnitude and durability of immune responses to Wildtype (WT) SARS-CoV-2 and Omicron VOCs, as compared to the initial 2-dose strategy. As we previously reported, relative to healthy controls, IMID patients exhibit accelerated waning of SARS-CoV-2 specific Ab, neutralization, and T cell responses after the second dose. The third dose restored and maximized responses to WT SARS-CoV-2 and broadened Ab responses against VOCs. Furthermore, the third and fourth doses enhanced the durability of immune responses post-vaccination compared to the second dose. We additionally demonstrate that SARS-CoV-2 specific T cell cytokine release increases with each successive booster vaccine. Finally, we show that anti-TNF treatment results in greater deficits in Ab responses in inflammatory bowel disease (IBD) patients compared to other anti-TNF-treated IMID patients. These results highlight the importance of third and fourth doses of mRNA vaccines in the immunocompromised to prolong and broaden responses to SARS-CoV-2 vaccination.

## Results

### Participant characteristics

161 participants contributed 607 samples over eight timepoints beginning in January 2021 (Figure 1, Table 1). Inclusion criteria were adult patients diagnosed with one or more immune-mediated inflammatory diseases (inflammatory bowel disease, rheumatoid arthritis, psoriatic arthritis, ankylosing spondylitis, psoriasis or hidradenitis suppurativa), untreated or treated with maintenance immunosuppressive therapy (anti-IL-12/23, anti-IL-17, anti-IL-23, anti-TNF, methotrexate/azathioprine (MTX/AZA), anti-TNF+MTX/AZA), and vaccinated with a SARS-CoV-2 mRNA vaccine (BNT162b2 Pfizer/BioNTech or mRNA-1273 Moderna). Henceforward, in our analyses, we denote patients treated with anti-TNF monotherapy or anti-TNF+MTX/AZA combination therapy as “TNF IMID” and IMID patients not treated with anti-TNF therapy as “non-TNF IMID.” The most common diagnosis was inflammatory bowel disease (IBD), and the most common treatments were anti-TNF-, anti-TNF+MTZ/AZA- and anti-IL-12/23-therapy. Healthy controls were included only up to 3 months post-dose 3. Most study subjects received the Pfizer vaccine. Age did not have significant effects on responses to vaccination among IMID patients (Supplemental Figure 1). There were no significant differences between vaccine intervals for dose 1-2 or dose 2-3 amongst treatment groups, with anti-TNF treated patients receiving slightly shorter intervals between 3^rd^ and 4^th^ dose compared to anti-IL-12/23- and anti-IL-23-treated patients (Supplemental Figure 2). Descriptive statistics of the samples used in the analyses of IgG, neutralization, and T cell cytokine and cytotoxic molecule data can be found in Supplemental Tables S1-S4.

**Figure 1.**
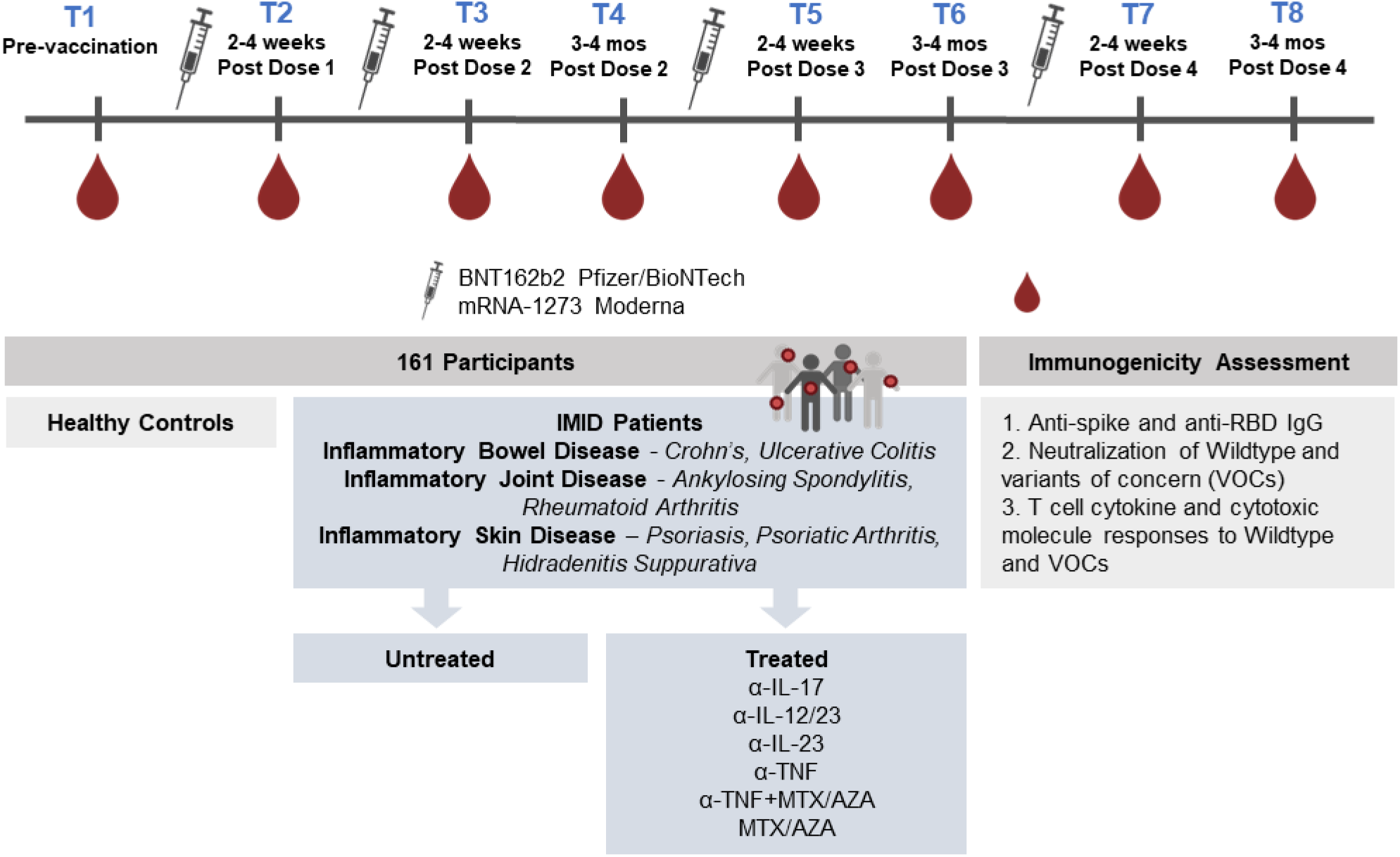
Schematic diagram of the IMPACT study. Blood was sampled from each patient at up to 8 timepoints spanning across pre-vaccination to after one to four SARS-CoV-2 mRNA vaccine doses for immunogenicity assessment. α-TNF, anti-tumor necrosis factor; MTX, methotrexate; AZA, azathioprine.

**Table 1.** Participant demographics.

|  | T1: Pre | T2: 2-4wks_PD1 | T3: 2-4wks_PD2 | T4: 3-4M_PD2 | T5: 2-4wks_PD3 | T6: 3-4M_PD3 | T7: 2-4wks_PD4 | T8: 3-4M_PD4 |
| --- | --- | --- | --- | --- | --- | --- | --- | --- |
| **IMID patients** |  |  |  |  |  |  |  |  |
| **Included in analysis - N** | 94 | 112 | 103 | 84 | 53 | 41 | 25 | 20 |
| Recruited - N | 96 | 119 | 112 | 90 | 57 | 49 | 32 | 25 |
| Excluded<sup>a</sup>- N | 2 | 7 | 9 | 6 | 4 | 8 | 7 | 5 |
| **Female - N (%)** | 55 (59) | 63 (56) | 58 (56) | 47 (56) | 28 (53) | 20 (49) | 14 (56) | 11 (55) |
| **Age - median (IQR)** | 39 (29 - 52) | 42.5 (30 - 55.5) | 43 (31 - 57) | 43 (30 - 56) | 44 (35 - 57) | 41 (30 - 49) | 58 (46 - 64) | 56 (36.5 - 65.5) |
| **BMI - median (IQR)** | 24 (22 - 27) | 24 (22 - 27) | 24 (22 - 27) | 24 (22 - 27) | 26 (23 - 31) | 25 (23 - 29) | 24 (22 - 29) | 24 (21 - 28) |
| **Diagnosis<sup>b</sup> - N (%)** |  |  |  |  |  |  |  |  |
| Psoriasis | 5 (5.3) | 16 (14.3) | 16 (15.5) | 18 (21.4) | 10 (18.9) | 5 (12.2) | 4 (16.0) | 4 (20.0) |
| Rheumatoid Arthritis | 2 (2.1) | 3 (2.7) | 4 (3.9) | 3 (3.6) | 2 (3.8) | 1 (2.4) | 2 (8.0) | 1 (5.0) |
| Ankylosing Spondylitis | 11 (11.7) | 11 (9.8) | 11 (10.7) | 3 (3.6) | 2 (3.8) | 0 (0.0) | 0 (0.0) | 0 (0.0) |
| Psoriatic Arthritis | 15 (16.0) | 20 (17.9) | 17 (16.5) | 13 (15.5) | 10 (18.9) | 8 (19.5) | 8 (32.0) | 5 (25.0) |
| Hidradenitis Suppurativa | 0 (0.0) | 0 (0.0) | 0 (0.0) | 0 (0.0) | 1 (1.9) | 1 (2.4) | 0 (0.0) | 0 (0.0) |
| Inflammatory Bowel Disease | 69 (73.4) | 71 (63.4) | 65 (63.1) | 56 (66.7) | 35 (66.0) | 28 (68.3) | 14 (56.0) | 12 (60.0) |
| Other | 6 (6.4) | 4 (3.6) | 2 (1.9) | 2 (2.4) | 0 (0.0) | 0 (0.0) | 1 (4.0) | 0 (0.0) |
| **Treatment group - N (%)** |  |  |  |  |  |  |  |  |
| Untreated | 9 (9.6) | 9 (8.0) | 9 (8.7) | 9 (10.7) | 5 (9.4) | 6 (14.6) | 0 (0.0) | 0 (0.0) |
| Anti-TNF | 37 (39.4) | 37 (33.0) | 34 (33.0) | 23 (27.4) | 16 (30.2) | 9 (22.0) | 5 (20.0) | 4 (20.0) |
| Anti-IL17A | 3 (3.2) | 6 (5.4) | 6 (5.8) | 6 (7.1) | 6 (11.3) | 5 (12.2) | 4 (16.0) | 3 (15.0) |
| MTX/AZA | 3 (3.2) | 7 (6.2) | 7 (6.8) | 4 (4.8) | 2 (3.8) | 1 (2.4) | 1 (4.0) | 2 (10.0) |
| Anti-TNF + MTX/AZA | 15 (16.0) | 16 (14.3) | 14 (13.6) | 10 (11.9) | 8 (15.1) | 6 (14.6) | 6 (24.0) | 5 (25.0) |
| Anti-IL12/23 | 26 (27.7) | 27 (24.1) | 23 (22.3) | 22 (26.2) | 11 (20.8) | 11 (26.8) | 7 (28.0) | 5 (25.0) |
| Anti-IL23 | 1 (1.1) | 10 (8.9) | 10 (9.7) | 10 (11.9) | 5 (9.4) | 3 (7.3) | 2 (8.0) | 1 (5.0) |
| **Days since vaccination - median (IQR)** |  | 26 (23 - 31) | 16 (14 - 19) | 104 (97 - 116) | 24 (18 - 34) | 110 (97 - 124) | 23 (19 - 28) | 100 (92 - 130) |
| **Vaccine type - N (%)** |  |  |  |  |  |  |  |  |
| Pfizer |  | 97 (86.6) | 78 (75.7) | 58 (69.0) | 32 (60.4) | 23 (56.1) | 15 (60.0) | 15 (75.0) |
| Moderna |  | 11 (9.8) | 7 (6.8) | 8 (9.5) | 3 (5.7) | 3 (7.3) | 0 (0.0) | 0 (0.0) |
| Mix of Pfizer & Moderna |  | 4 (3.6) | 18 (17.5) | 18 (21.4) | 18 (34.0) | 15 (36.6) | 10 (40.0) | 5 (25.0) |

| Healthy controls |  |  |  |  |  |  |
| --- | --- | --- | --- | --- | --- | --- |
| **Included in analysis - N** | 15 | 17 | 21 | 11 | 5 | 6 |
| Recruited - N | 16 | 18 | 22 | 12 | 6 | 6 |
| Excluded<sup>a</sup> - N | 1 | 1 | 1 | 1 | 1 | 0 |
| **Female - N (%)** | 7 (47) | 7 (41) | 8 (38) | 4 (36) | 3 (60) | 3 (50) |
| **Age - median (IQR)** | 33 (25 - 46) | 34 (26 - 42) | 34 (26 - 46) | 27 (25 - 47) | 27 (25 - 37) | 31 (26 - 37) |
| **BMI - median (IQR)** | 24 (23 - 27) | 25 (23 - 27) | 25 (22 - 28) | 23 (22 - 27) | 23 (23 - 28) | 24 (22 - 27) |
| **Days since vaccination – median (IQR)** |  | 23 (21 - 27) | 16 (13 - 19) | 101 (95 - 105) | 21 (15 - 30) | 94 (92 - 99) |
| **Vaccine type - N (%)** |  |  |  |  |  |  |
| Pfizer |  | 16 (94.1) | 19 (90.5) | 9 (81.8) | 4 (80.0) | 4 (66.7) |
| Moderna |  | 0 (0.0) | 0 (0.0) | 0 (0.0) | 0 (0.0) | 0 (0.0) |
| Mix of Pfizer & Moderna |  | 1 (5.9) | 2 (9.5) | 2 (18.2) | 1 (20.0) | 2 (33.3) |

<sup>a</sup> Excluded from analysis due to evidence of SARS-CoV-2 infection or missing covariates used in analysis (i.e., BMI [n=13] or vaccine type [n=1])
<sup>b</sup> Diagnosis are not mutually exclusive. Participants may have reported multiple diagnoses
Abbreviations: AZA: azathioprine; BMI: Body Mass Index; IL: Interleukin; IMID: Immune-Mediated Inflammatory Diseases; IQR: Inter-quartile range; M: months; MTX: Methotrexate; PD: Post dose; TNF: Tumor necrosis factor; wks: weeks

### Reduced humoral and cellular responses in IMID patients

To compare humoral and cellular responses post-vaccination in untreated and treated IMID patients to healthy controls, linear regression models (controlling for age, BMI, sex, and vaccine type) were generated at each timepoint following the first, second and third vaccine dose (T2-T6, as defined in Figure 1 and Table 1). The predicted mean difference between each study group relative to healthy controls is plotted in Figure 2. Anti-TNF-, anti-TNF+MTX/AZA-, and anti-IL-23-treated patients displayed reduced spike- or RBD-specific Ab levels relative to healthy controls 2-4 weeks after the first dose of vaccine (T2); however, most deficits were corrected by a second dose (T3) (Figure 2A). By 3-4 months after the second dose (T4), anti-TNF- and anti-TNF+MTX/AZA-treated patients displayed significantly reduced SARS-CoV-2-specific Ab responses, while all other IMID patients maintained comparable levels to healthy controls (Figure 2A). All IMID patients tended to have a reduced capacity to neutralize WT SARS-CoV-2 compared to healthy controls after dose 2 (T4) in a spike-pseudotyped lentiviral assay (Figure 2A). A third dose of vaccine (T5) corrected deficits in S- and RBD-specific Ab levels and neutralization responses observed in IMID patients. 3-4 months after dose 3 (T6), only anti-TNF- and anti-TNF+MTX/AZA-treated patients displayed reduced antigen (Ag)-specific Ab and neutralization responses compared to healthy controls.

**Figure 2.**
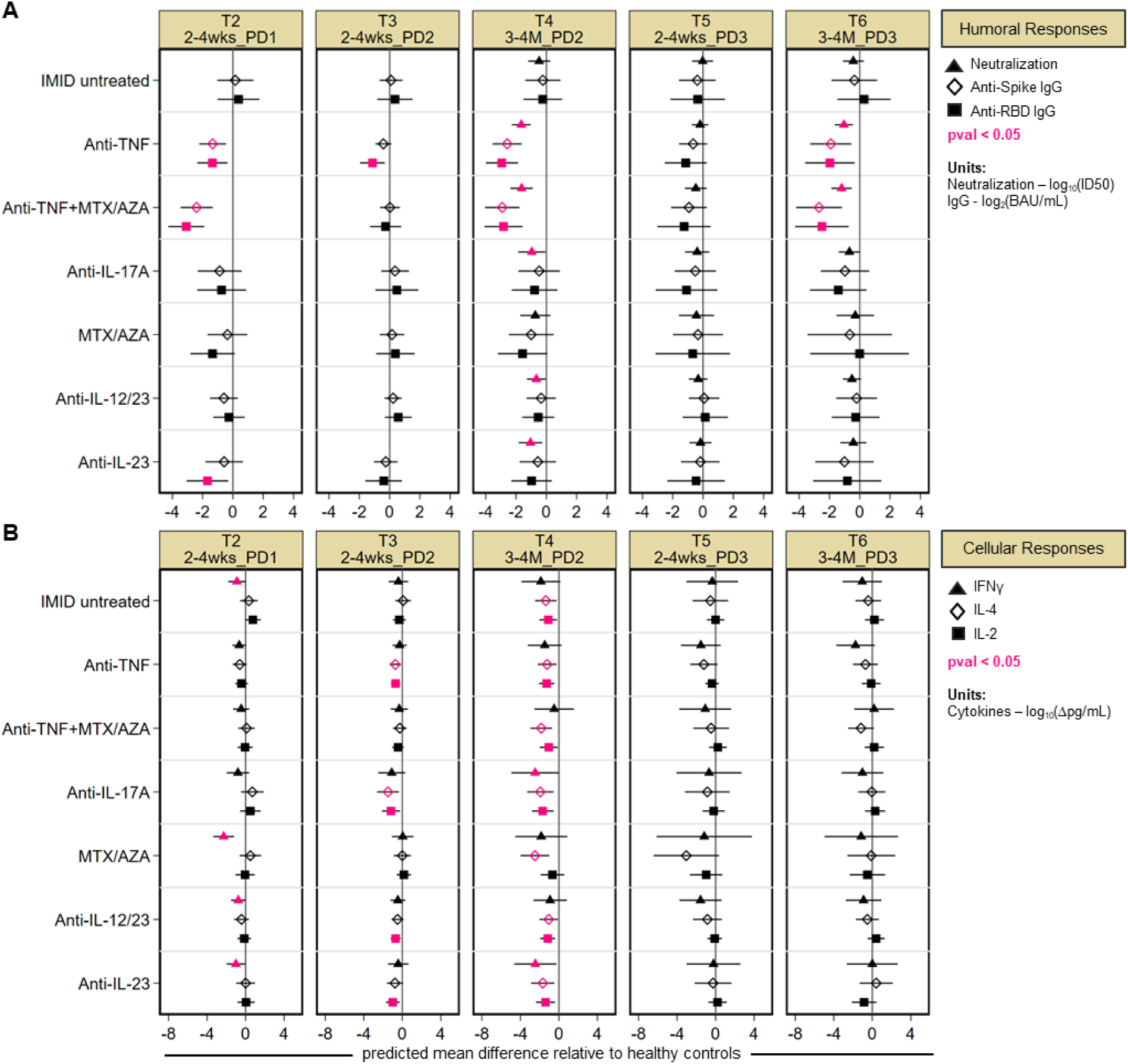
Predicted mean difference in humoral and cellular responses in treated IMID patients relative to healthy controls, after one to three vaccine doses. Patients are stratified by treatment group. Timepoints are defined above each panel, as described in Table 1. **(A)** Mean difference in neutralization against Wildtype (WT) SARS-CoV-2 (triangle), anti-spike and anti-RBD IgG (diamond and square respectively) **(B)** Mean difference in T cell IFNγ (triangle), IL-4 (diamond), IL-2 (square) responses after vaccination in IMID patients. **(A, B)** Least-squares linear regression models controlled for age, BMI, sex, and vaccine type. Significant mean differences (p-value < 0.05) are colored in pink. 95% confidence intervals are plotted across each point. **(A-C)** For neutralization, IgG, and cytokine data, sample sizes are listed in Supplemental Table 1. log_10_(ID50), the serum dilution that inhibits 50% of lentivirus infection; BAU/mL, binding antibody units per milliliter; Δ, background subtracted; pg/mL, picogram per milliliter; PD: post dose

With regards to memory T cell responses to SARS-CoV-2, after one dose of vaccine (T2), IMID untreated, MTX/AZA-, anti-IL-12/23-, and anti-IL-23-treated patients had reduced IFNγ production relative to healthy controls, and these deficits were corrected by a second dose (T3) (Figure 2B). IL-2 and/or IL-4 production was reduced in several treated IMID groups after dose 2 (T3). 3-4 months later (T4), all IMID patients displayed significantly reduced IL-2 and IL-4 production relative to healthy controls (Figure 2B). Reduced IFNγ production was also observed in anti-IL-17A- and anti-IL-23-treated patients at T4 (Figure 2B). No deficits in cytokine responses relative to healthy controls were observed after the third dose (T5, T6) (Figure 2B).

In sum, IMID patients had significantly reduced SARS-CoV-2 specific Ab, neutralization, and T cell responses to SARS-CoV-2 3-4 months after dose 2. A third vaccine dose was critical for boosting these responses to levels similar to healthy controls. The greatest deficits were observed in the SARS-CoV-2 specific Ab responses of anti-TNF- and anti-TNF+MTX/AZA-treated patients. Therefore, in subsequent analyses, we included comparisons between TNF IMID and non-TNF IMID patients.

### Reduced antibody responses in anti-TNF treated subjects across four vaccine doses

To parse the effects of the third and fourth vaccine doses on the magnitude of Ab responses, mixed-effects linear regression models (controlling for age, BMI, and sex, with an interaction term between timepoints and study group) were generated to predict the mean anti-RBD and anti-spike IgG responses at each timepoint (Figure 3). First and second vaccine doses significantly increased anti-RBD and anti-spike IgG levels compared to baseline, with maximal levels achieved after 2 vaccine doses in all participants (Figure 3A-D, Supplemental Table S5). As noted in the pairwise analysis in Figure 1, this longitudinal analysis confirms that by 3-4 months post dose 2 (T4), IMID patients exhibited reduced anti-RBD and anti-spike levels compared to healthy controls (Figure 3A-B). This was largely driven by TNF IMID patients (Figure 3C-D). The third and fourth vaccine doses (T5-T8) did not result in significant measurable increases in Ag-specific Ab levels over the peak dose 2 response, with no overall differences between healthy controls and IMID patients (Figure 3A-D). A limitation to this conclusion is that many healthy controls and non-TNF IMID patients reached the upper limit of quantification for anti-spike IgG levels after the second dose of vaccine (T3), with the percentage of patients reaching saturating Ab levels increasing with subsequent doses (Supplemental Table S3). This was less of an issue for RBD-specific IgG responses as levels were generally lower. Although IMID patients as a whole reached the same peak Ab responses as healthy controls, TNF IMID patients consistently showed reduced RBD- and spike-specific Ab levels compared to healthy controls or non-TNF IMID patients across 1 to 4 doses of vaccine (Figure 3C-D).

**Figure 3.**
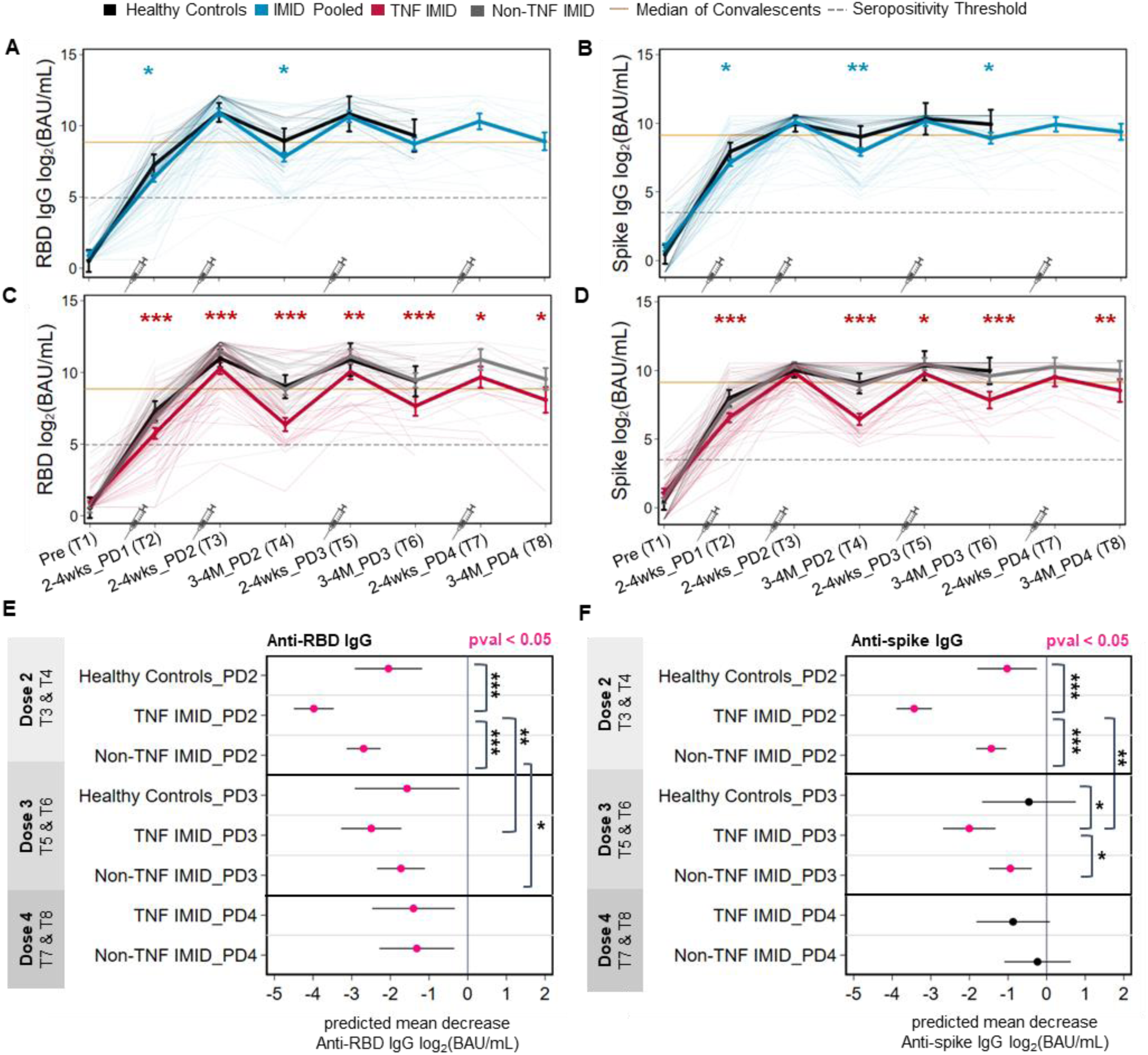
Longitudinal analyses of the magnitude and durability of antibody responses pre and post one to four doses of SARS-CoV-2 mRNA vaccine. Timepoints are defined in Table 1. Predicted mean anti-RBD IgG **(A, C)** and anti-spike IgG **(B, D)** levels log_2_(BAU/mL)) from pre-vaccination to post one to four doses of vaccine in healthy controls and IMID patients. **(A-D)** Mixed-effects linear regression models controlled for age, BMI, and sex (thick lines), with an interaction term between timepoint and study group. Individual patients are plotted in thin lines. Study groups: healthy controls (black), IMID patients pooled (blue), TNF IMID (anti-TNF or anti-TNF+MTX/AZA treated) patients (red), and IMID patients not treated with anti-TNF (grey). The dashed grey lines represent the IgG seropositivity threshold. The yellow lines represent the median IgG levels from 340 convalescent cases (PCR-confirmed COVID-19 cases 21–115 days after symptom onset). An additional control for vaccine type was incorporated into mixed-effects regression models to compare groups: IMID pooled vs healthy controls (blue asterisks) or TNF IMID vs IMID pooled (red asterisks). **(E, F)** Predicted mean decrease of anti-RBD IgG **(E)** and anti-spike IgG **(F)** in healthy controls, TNF IMID, and IMID patients not treated with anti-TNF (between 2-4 weeks and 3-4 months) after dose 2 (decrease between T3 and T4), after dose 3 (decrease between T5 and T6), and after dose 4 (decrease between T7 and T8). Mixed-effects linear regression models controlled for age, BMI, sex, vaccine type, with an interaction between timepoint and study group. Values that are significant (pval<0.05) are colored in pink. Significant pairwise comparisons are indicated by asterisks and brackets. **(A-F)** Sample sizes are listed in Supplemental Table 1. *p<0.05; **p<0.01; ***p<0.001; BAU/mL, binding antibody units per milliliter; PD, post dose

### Third and fourth doses enhance the durability of IgG responses in IMID patients

While the third and fourth vaccine dose had little or no effects on the overall magnitude of Ab responses, booster doses were critical for limiting the decay of Ab levels post-vaccination. Using mixed-effects linear regression models, we predicted the mean decrease in RBD- and spike-specific IgG levels between 2 timepoints (2-3 weeks and 3-4 months) after doses 2, 3, and 4 (Figure 3E-F). All groups exhibited significant decreases in anti-RBD and anti-spike levels by 3- 4 months after dose 2, with the greatest decreases observed in TNF IMID patients. The degree of waning between anti-RBD and anti-spike IgG following the third and fourth vaccine doses showed distinct kinetics. For anti-RBD IgG, all study groups showed waning of responses at 3-4 months after dose 3, albeit of reduced magnitude relative to that observed after the second dose, whereas there were no differences in waning between the third and fourth doses (Figure 3E). Anti-spike IgG levels did not significantly decline in healthy controls by 3-4 months after dose 3, whereas IMID patients’ spike-specific IgG levels still waned after dose 3 (Figure 3F). The fourth dose maintained spike-specific IgG levels such that decreases observed in IMID patients in the 3-4 months following the fourth vaccination dose were no longer significant (Figure 3F). In sum, the third and fourth doses of vaccine in IMID participants enhanced the duration of Ab responses to SARS-CoV-2.

### Effect of third and fourth vaccine doses on neutralization activity against Wildtype SARS-CoV-2 and VOCs

Our previous work demonstrated that after 2 vaccine doses, IMID patients had a reduced ability to neutralize WT SARS-CoV-2 compared to healthy controls and TNF IMID patients could not neutralize the Omicron BA.1 VOC (20). Here we used mixed-effects linear regression models (controlling for age, BMI, sex, and vaccine type, with an interaction term between timepoint and study group) to predict neutralization activity following 2 to 4 vaccine doses (Figure 4).

**Figure 4.**
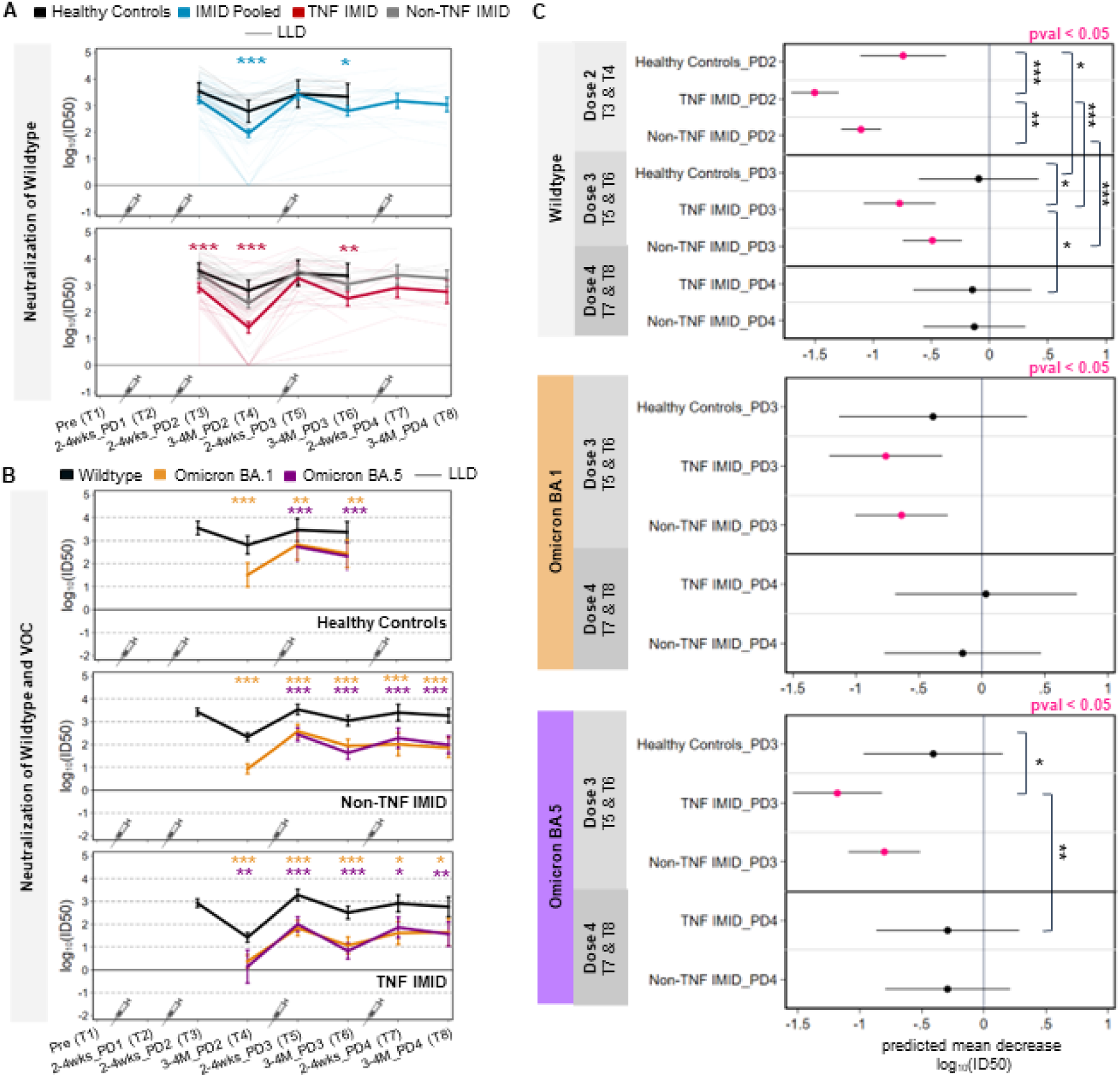
Longitudinal analyses of the magnitude and durability of neutralization responses post two to four doses of SARS-CoV-2 mRNA vaccine. Timepoints are defined in Table 1. **(A)** Predicted neutralization (log_10_(ID50)) activity against Wildtype (WT) SARS-CoV-2 after two to four doses of vaccine, grouped by healthy controls (black), IMID patients pooled (blue), TNF IMID (anti-TNF or anti-TNF+MTX/AZA treated) patients (red), and IMID patients not treated with anti-TNF (grey). Comparisons between groups are indicated: IMID pooled vs healthy controls (blue asterisks) or TNF IMID vs IMID pooled (red asterisks). **(B)** Predicted neutralization (log_10_(ID50)) activity against WT, Omicron BA.1 and BA.5 after two to four doses of vaccine, grouped by healthy controls, IMID patients not treated with anti-TNF and TNF IMID patients. Neutralization activity against WT is depicted in black, against Omicron BA.1 in orange, and against Omicron BA.5 in purple. Paired T-tests were conducted to compare responses to Omicron BA.1/BA.5 versus WT. **(C)** Predicted mean decrease of neutralization against WT and Omicron BA.1/BA.5 in healthy controls, TNF IMID, and IMID patients not treated with anti-TNF (between 2-4 weeks and 3-4 months) after dose 2 (decrease between T3 and T4), after dose 3 (decrease between T5 and T6), and after dose 4 (decrease between T7 and T8). Mean decrease values that are significant (pval<0.05) are colored in pink. Significant pairwise comparisons are indicated by asterisks and brackets. **(A, C)** Sample sizes are listed in Supplemental Table 1. **(B)** Sample sizes are listed in Supplemental Table 2. **(A-C)** Mixed-effects linear regression models controlled for age, BMI, sex, vaccine type, with an interaction term between timepoint and study group. *p<0.05; **p<0.01; ***p<0.001; log_10_(ID50), the serum dilution that inhibits 50% of lentivirus infection; BAU/ml, binding antibody units per milliliter; LLD, lower limit of detection; PD: post dose

3-4 months after dose 2, 10% of IMID patients had no detectable neutralizing activity (lower limit of detection) against WT SARS-CoV-2, and the IMID group overall had significantly reduced neutralization activity compared to healthy controls (Figure 4A). This difference between healthy controls and IMID patients was corrected by the third dose. While peak neutralization levels were reached in healthy donors after 2 doses of vaccine, for IMID patients, the third dose increased neutralization levels from dose 2 (p=0.019), driven by an increase in the TNF IMID patients (p=0.005), with no further enhancement by the fourth dose (Supplemental Table S5).

Neutralization activity against BA.1 and BA.5 VOCs was significantly weaker than neutralization against WT at all timepoints for all study groups, where data were available (Figure 4B, all p<0.05). The majority of TNF IMID patients were unable to neutralize either Omicron BA.1 or Omicron BA.5 3-4 months after dose 2 but developed neutralization activity after the third dose (Figure 4B). At each timepoint, a significant proportion of TNF IMID patients had neutralization responses to BA.1 and BA.5 that were at the lower limit of detection (Supplemental Table S4); hence the deficits in their responses to VOCs relative to the WT may be underestimated.

While the third and fourth doses minimally affected the magnitude of neutralization activity against Wildtype and VOCs, compared to the peak response after the second dose, the boosters were critical for prolonging responses. Between 2-4 weeks and 3-4 months after dose 2, all participants displayed significant decreases in neutralization activity against WT SARS-CoV-2, with TNF IMID patients displaying the largest decrease (Figure 4C). Neutralization levels against WT SARS-CoV-2 and VOCs did not significantly decline in healthy donors by 3 months after the third dose, whereas IMID patients continued to show waning in neutralization responses after dose 3 (Figure 4C). However, 3 months after dose 4, there was no longer a significant decay in neutralization responses to WT or VOCs among all IMID patients (TNF IMID and non-TNF IMID) (Figure 4C).

Together, these data suggest that third doses are important in healthy controls and IMID patients for reducing the decay of neutralization responses to all variants of SARS-CoV-2, thereby broadening neutralization activity, with fourth doses showing additional stabilization effects in IMID patients.

### Anti-TNF therapy significantly impairs humoral responses to SARS-CoV-2 in IBD patients

Given the observation that anti-TNF therapy had the most profound effect on Ab responses to the SARS-CoV-2 vaccine, we next investigated whether the specific disease type impacted these results. Based on the sample size, this analysis was limited to the stratification of IMID patients by IBD diagnosis. Ab responses post-dose 2 (T3 and T4), post-dose 3 (T5 and T6), and post-dose 4 (T7 and T8) were pooled. Regardless of vaccine dose, anti-spike IgG and anti-RBD IgG were significantly lower for IBD patients treated with anti-TNF therapy compared to those who were not treated with anti-TNF therapy (Figure 5A-F). At timepoints after doses 2 and 3, neutralization capacity was lower for IBD patients on anti-TNF therapy compared to IBD patients not treated with anti-TNF (Figure 5G-H). In contrast, treatment status had no significant impact on Ab responses among IMID patients who did not report an IBD diagnosis (Figure 5).

**Figure 5.**
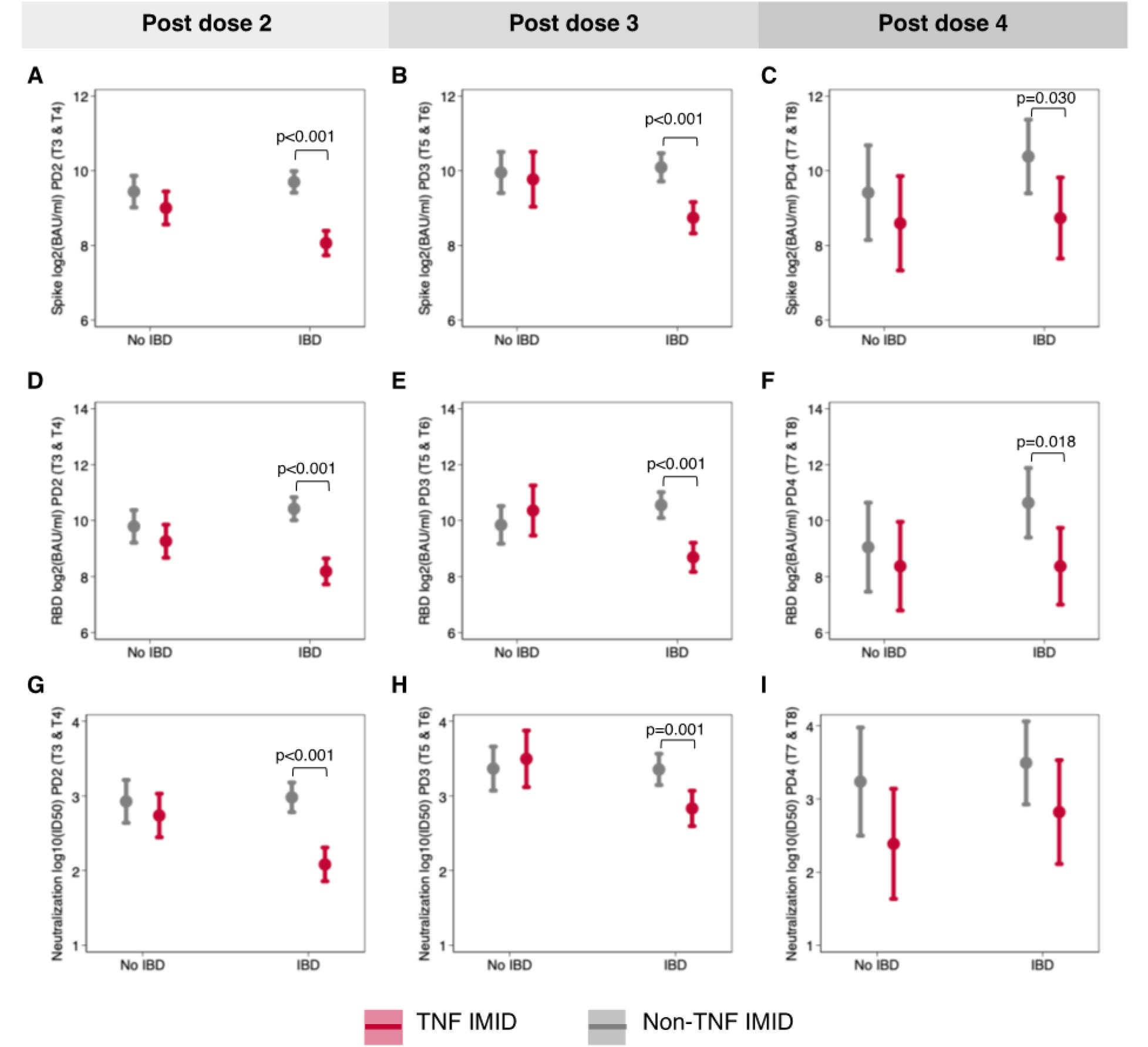
Impact of anti-TNF therapy by disease type. The effect of anti-TNF therapy on anti-spike IgG **(A-C)** anti-RBD IgG **(D-F)** and neutralization of Wildtype SARS-CoV-2 **(G-I)** was tested for participants with and without inflammatory bowel disease (IBD) using mixed effects models with an interaction term between IBD status and treatment with anti-TNF therapy. Models controlled for days post vaccination, sex, age, BMI, and vaccine type, and were repeated for post dose 2 **(A, D, G)**, post dose 3 **(B, E, H)**, and post dose 4 **(C, F, I)** timepoints. For IgG data, samples sizes for each sub-group were as follows: N = 38 for non-IBD/non-TNF/PD2, N = 28 for non-IBD/TNF/PD2, N = 68 for IBD/non-TNF/PD2, N = 58 for IBD/TNF/PD2, N = 21 for non-IBD/non-TNF/PD3, N = 9 for non-IBD/TNF/PD3, N = 34 for IBD/non-TNF/PD3, N = 29 for IBD/TNF/PD3, N = 10 for non-IBD/non-TNF/PD4, N = 9 for non-IBD/TNF/PD4, N = 44 IBD/non-TNF/PD4, and N = 11 for IBD/TNF/PD4. For neutralization data, samples sizes for each sub-group were as follows: N = 36 for non-IBD/non-TNF/PD2, N = 28 for non-IBD/TNF/PD2, N = 67 for IBD/non-TNF/PD2, N = 51 for IBD/TNF/PD2, N = 20 for non-IBD/non-TNF/PD3, N = 9 for non-IBD/TNF/PD3, N = 29 for IBD/non-TNF/PD3, N = 25 for IBD/TNF/PD3, N = 8 for non-IBD/non-TNF/PD4, N = 8 for non-IBD/TNF/PD4, N = 11 for IBD/non-TNF/PD4, N = 6 for IBD/TNF/PD4.

For each outcome, at timepoints post doses 2 and 3, there was a significant interaction between IBD status and anti-TNF treatment, such that the effect of anti-TNF therapy was significantly greater for IBD patients than for other IMID patients (Figure 5A-B, 5D-E, 5G-H). In these data, anti-TNF monotherapy treated groups, as well as patients treated with anti-TNF+MTX/AZA combination therapy, were pooled into the anti-TNF treated category (TNF IMID). However, the removal of the combination therapy groups did not affect the significance of these results.

### Third and fourth doses increase the magnitude and durability of T cell cytokine responses

To elucidate the effects of the third and fourth doses of vaccine on the magnitude and durability of T cell responses post vaccination, mixed-effects linear regression models (controlling for age, sex, and BMI, with an interaction term between timepoint and study group) were used to estimate the mean levels of secreted T cell cytokine IL-2, IL-4, IFNγ (Figure 6), as well as IL-17 and cytotoxic molecules (Supplemental Figure 3) responses across 1 to 4 doses of vaccine.

**Figure 6.**
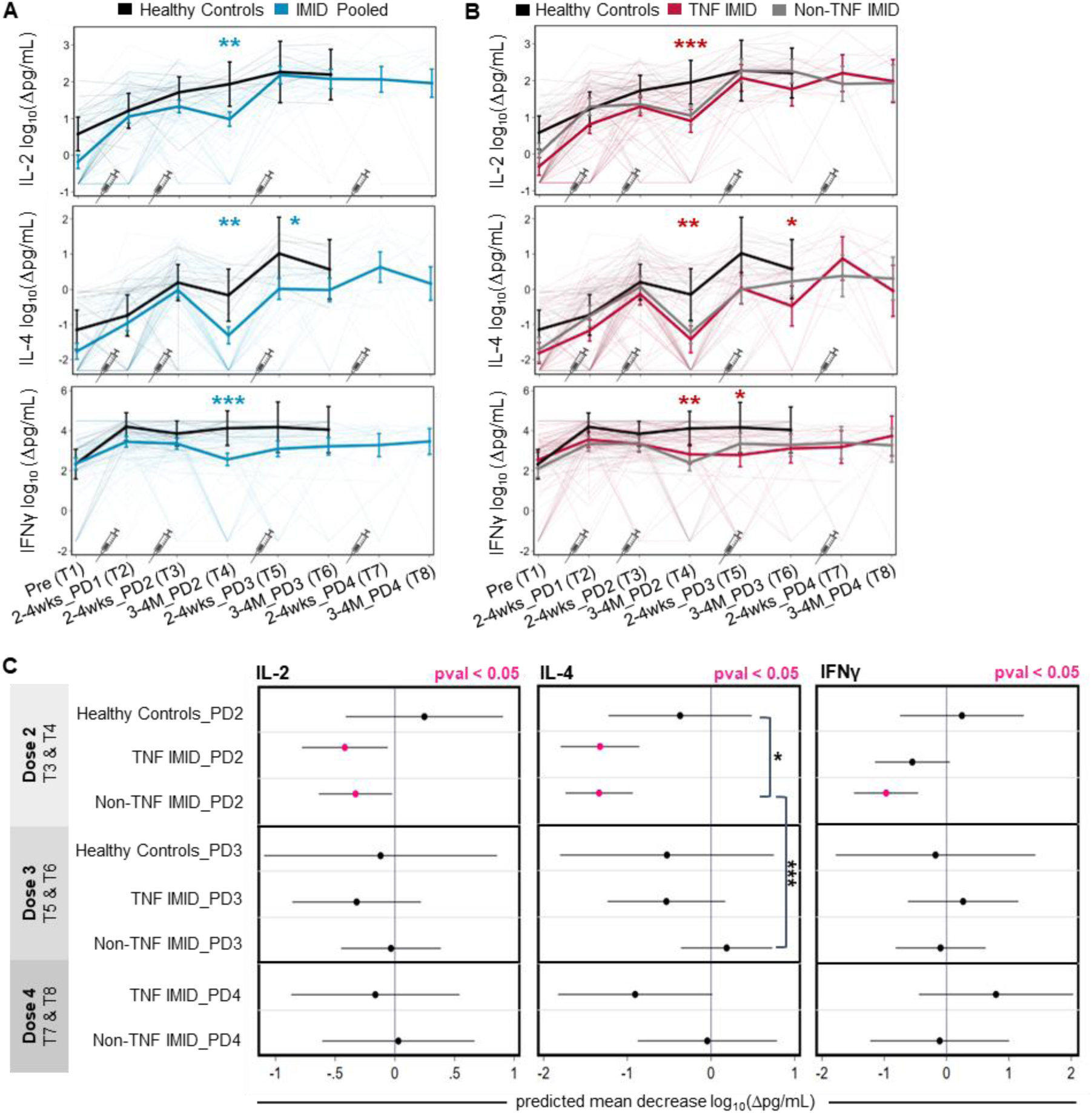
Longitudinal analyses of T cell cytokine responses pre and post one to four doses of SARS-CoV-2 mRNA vaccine. Timepoints are defined in Table 1. **(A, B)** Predicted mean T cell cytokines IL-2, IL-4, IFNγ (log_10_ (Δpg/mL)) from pre-vaccination to after one to four doses of vaccine. Mixed-effects linear regression models controlled for age, BMI, and sex, with an interaction term between timepoint and group (thick lines). Individual patients are plotted in thin lines. Groupings: healthy controls (black), IMID patients pooled (blue), TNF IMID (anti-TNF or anti-TNF+MTX/AZA treated) patients (red), and IMID patients not treated with anti-TNF (grey). An additional control for vaccine type was incorporated into mixed-effects regression models to compare groups: a) IMID pooled vs healthy controls (blue asterisks) or b) TNF IMID vs healthy controls (red asterisks)**. (C)** Predicted mean decrease of T cell cytokines IL-2, IL-4, IFNγ (log_10_ (Δpg/mL)) in healthy controls, TNF IMID, and IMID patients not treated with anti-TNF between 2-4 weeks and 3-4 months after dose 2 (decrease between T3 and T4), after dose 3 (decrease between T5 and T6), and after dose 4 (decrease between T7 and T8). Mixed-effects linear regression models controlled for age, BMI, sex, vaccine type, with an interaction between timepoint and group. Mean decrease values that are significant (pval<0.05) are colored in pink. Significant pairwise comparisons are indicated by asterisks and brackets. **(A-C)** Sample sizes are listed in Supplemental Table 1. *p<0.05; **p<0.01; ***p<0.001; Δ: background subtracted; pg/mL: picogram per milliliter; PD: post dose

Relative to baseline (T1), the first and second doses of vaccine (T2, T3) boosted IL-2, IL-4, and IFNγ production in healthy controls and IMID patients (Figure 6A). Peak IL-2 responses were observed by the third dose in healthy controls and IMID patients, with no differences evident between the groups after dose 3 and no further impact of dose 4. T cell IL-4 responses increased with each successive dose, measured up to dose 3 in healthy donors and dose 4 in IMID patients (Figure 6A, Supplemental Table S5). Notably, there was a significant increase in IL-4 between dose 3 and dose 4 for TNF IMID patients (p=0.026), which was not apparent in non-TNF IMID patients (Figure 6B). This effect was primarily driven by patients on combination therapy with anti-TNF plus MTX/AZA (data not shown). IFNγ production peaked after the first dose for all participants (Figure 6A-B). After dose 2, IMID patients exhibited reductions in IFNγ production compared to healthy controls and the third dose restored but did not further boost IFNγ responses (Figure 6A-B). The fourth dose did not increase the magnitude of IFNγ responses beyond the maximum achieved after the first dose. TNF IMID patients exhibited similar IL-2, IL-4, and IFNγ responses to non-TNF IMID patients, with waning of cytokines after dose 2 of vaccine relative to healthy controls, and restoration of responses with a third and fourth dose (Figure 6B).

Other molecules assessed included IL-17A and cytotoxic molecules Granzyme A, Granzyme, Perforin, and sFasL. In all cases, production peaked after one dose of vaccine. Analogous to the IL-2, IL-4, and IFNγ responses, IMID patients exhibited reduced levels of cytotoxic molecules compared to healthy controls at 3-4 months after dose 2 (Supplemental Figure 3A-B), with these lower levels corrected by the third or fourth dose of vaccine.

To assess the effects of the third and fourth doses on prolonging T cell cytokine and cytotoxic molecule responses post-vaccination, we compared the predicted decrease in responses between 2 timepoints (2-4 weeks and 3-4 months) after the second, third and fourth doses of vaccination, respectively (Figure 6C). While healthy donor T cell responses were stable out to 3 months after dose 2, TNF IMID patients and non-TNF IMID patients exhibited significant decreases in IL-2 and IL-4 secretion after the second dose, with IL-4 exhibiting the most substantial decrease (Figure 6C). Non-TNF IMID patients also exhibited decreases in IFNγ responses after dose 2 (Figure 6C). However, after the third or fourth doses, there was no longer any significant decay in T cell cytokine responses over the next 3 months, highlighting the enhanced durability of cytokine responses in IMID patients after booster doses of vaccine. Parallel phenomena were observed with IL-17A, Granzyme A, Granzyme B, sFasL, and Perforin responses; all IMID patients, but not healthy controls, displayed significant decreases in responses after dose 2, with no significant decreases after booster doses (Supplemental Figure 4).

We additionally assessed T cell responses to BA.1 and BA.4/5 VOCs after the third dose of vaccine (T5, T6). Healthy controls and IMID patients (pooled) did not exhibit significant differences in IL-2, IL-4, IFNγ, sFasL, and Granzyme A responses to Omicron BA.1 or BA.4/5 compared to WT (Supplemental Figure 5, 6). Reduced IL-17A responses to BA.1 and BA.4/5 were observed at T6, and minor reductions in Granzyme B and Perforin responses to BA.1 or BA.4/5 were observed at T5 and T6 (Supplemental Figure 5, 6). Altogether, these data show that after 3 vaccine doses, T cell responses of IMID patients to WT and the BA.1 and BA.5 VOCs are largely equivalent.

### Correlations of T cell cytokine responses with neutralization responses post two and three doses of vaccination

Linear regression models were used to estimate the association between T cell and neutralization responses, focusing on IL-2 and IL-4 responses in IMID patients. Of note, IL-2 responses after dose 2 significantly predicted neutralization 3-4 months later in both TNF IMID (p=0.019) and non-TNF IMID (p=0.017) patients (Figure 7A). This effect was no longer significant after dose 2, perhaps suggesting saturation of the T cell IL-2 response with respect to help for Ab responses (Figure 7B). There was also a significant correlation between IL-4 production at 2-4 weeks after dose 2 and neutralizing capacity 3-4 months later, among non-TNF IMID patients (p=0.001) but not among TNF IMID patients (Figure 7C). Following dose 3, the effect of IL-4 was no longer predictive of neutralizing capacity among the non-TNF IMID patients but became predictive in TNF IMID patients (p=0.008; Figure 7D). Accordingly, there is a significant difference in the effect of IL-4 on neutralizing capacity between study groups (p=0.015; Figure 7D). These data suggest that the early T cell responses may be important in defining the duration of the Ab response, with the timing of the impact of T cells related to when the maximal T cell signal is achieved.

**Figure 7.**
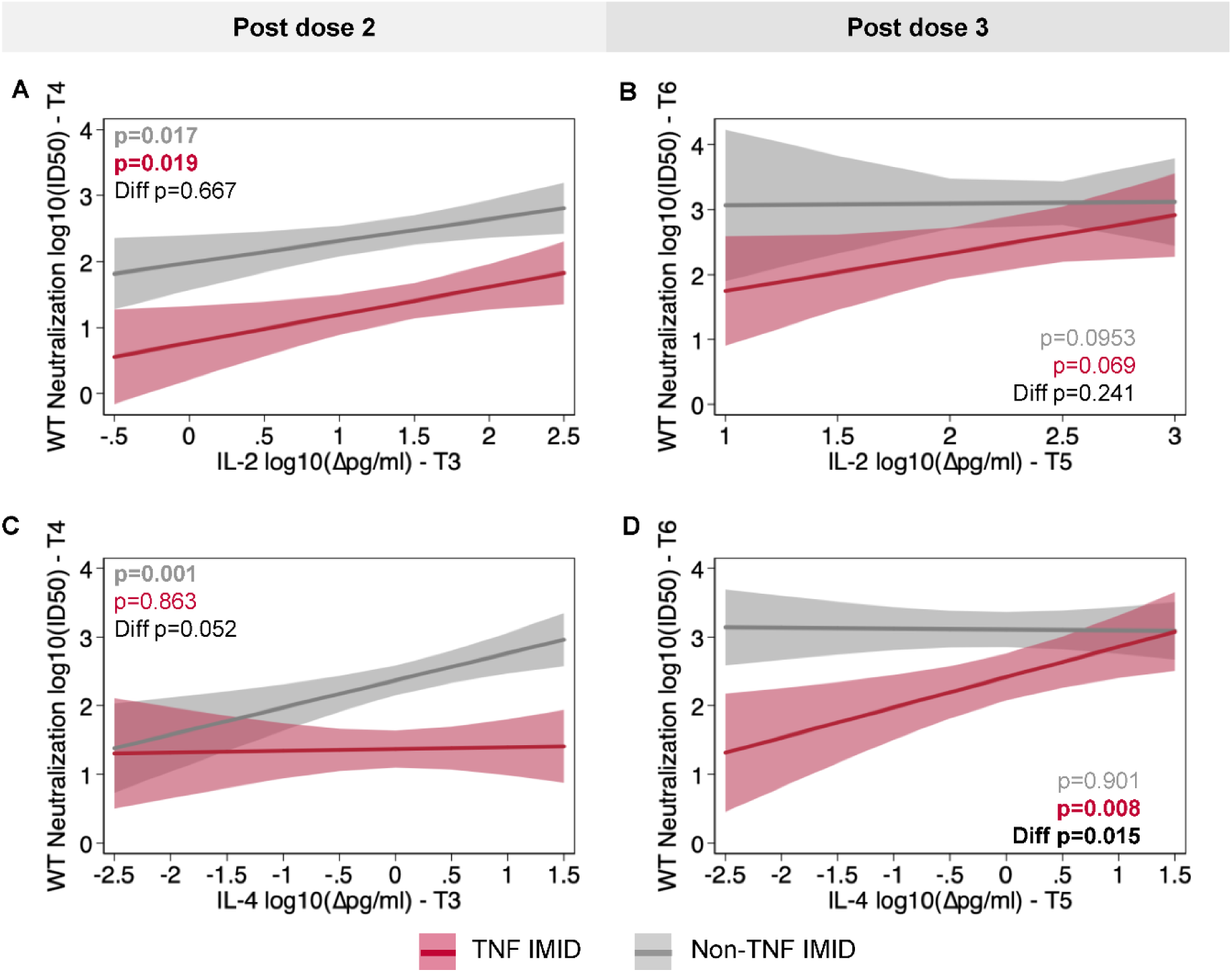
Association between T cell responses 2-4 weeks post-vaccination and neutralizing antibody titers 3 months later. The association of IL-2 **(A-B)** or IL-4 **(C-D) s**hortly after vaccination (T3: 2-4wks_PD2 or T5: 2-4wks_PD3) with neutralization capacity of the wild type strain of SARS-CoV-2 3 months later (T4: 3-4M_PD3 or T6: 3-4M_PD4) was tested using linear regression models that controlled for age, BMI, sex, and vaccine type. Regression models included an interaction term between cytokine levels and study group (i.e., Non-TNF IMID and TNF IMID) and were repeated post dose 2 **(A, C)** and post dose 3 **(B, D).**

## Discussion

In this longitudinal study, we followed a cohort of patients diagnosed with one or more immune-mediated inflammatory diseases to assess their responses to SARS-CoV-2 after one through four vaccine doses. We and others have reported that specific immunosuppressants significantly attenuate the magnitude and stability of humoral and cellular responses after 2 doses of vaccine in IMID patients compared to healthy controls, with the most pronounced deficits in TNF IMID patients (7, 10, 20, 24).

While T cell cytokine responses increased with successive vaccine doses, we observed no difference in the magnitude of the peak Ab response following third or fourth doses in healthy donors and IMID patients. There is debate in literature regarding the effect of a third dose on the serological responses in IMID patients. In studies in which patients have absent or weak serological responses after the second dose, a third dose significantly boosted Ab levels (11, 25, 26), while some IMID patients do not respond even after a third dose (12). It has also been observed that a third dose does not significantly boost levels compared to the peak after the second dose, which may be attributed to the stronger response seen in these individuals after the second dose as compared to weak responders (8). While the third and fourth doses had little or no measurable effects on the magnitude of Ab responses, the third dose, and to a lesser extent the fourth dose, was critical in enhancing the durability of Ab responses in the ensuing 3 months post-vaccination in healthy controls and IMID patients. These data are consistent with other studies in IMID and healthy patients demonstrating a slower decline in Ab levels after the third dose of vaccine (11, 27, 28). It is important to note that the immunogenicity of a third and fourth dose may differ based on the strength of response after the primary series of vaccination and the degree of immunosuppression, disease indication, type of treatment, and variability in assay dynamic ranges.

Irrespective of 1 to 4 doses of vaccine, anti-TNF treated patients consistently showed reduced Ab levels compared to healthy controls and non-TNF IMID patients, suggesting a long-term impact of anti-TNF treatment on the humoral response to vaccines. These results are consistent with another recent study (29). In contrast, T cell responses in anti-TNF and non-TNF IMID patients were similar. Strikingly, the negative effect of anti-TNF therapy on humoral responses was only observed in IBD patients. Removal of the combination-treated anti-TNF patients from this analysis did not affect the differences observed. This finding is in line with literature reporting an effect of anti-TNF therapy on SARS-CoV-2 vaccine response in IBD patients (10, 24, 30–32), but not in other IMID patients without an IBD diagnosis (33, 34). The mechanisms underlying the differential effect of anti-TNF therapy on vaccine-induced immunity based on disease indication are unclear but may be attributed to higher doses or a higher proportion receiving drugs intravenously in the IBD group compared to the non-IBD IMID patients, although this remains to be determined. Regardless of the mechanism, IBD patients on anti-TNF therapy have impaired humoral responses to vaccination and thus may be more susceptible to SARS-CoV-2 irrespective of vaccination status. These data support early booster vaccination regimens for these patients.

Additionally, the third dose of vaccine was necessary for IMID patients to develop robust neutralization activity against BA.1 and BA.5 VOCs, which was lacking in anti-TNF treated IMID patients 3-4 months after dose 2 (20), thereby broadening immune responses against SARS-COV-2. The kinetics of neutralization responses against WT and VOCs varied between healthy and IMID patients. While healthy controls had stable neutralization responses to WT and VOCs out to 3 months post vaccination after a third dose, IMID patients required a fourth dose to achieve this effect. Irrespective of the study group, neutralization activity against BA.1 and BA.5 VOCs was significantly weaker than neutralization of WT across 2 to 4 doses of vaccine, in line with studies demonstrating reduced geometric mean titers of neutralization against Omicron sub-lineages compared to the original Wildtype strain after 3 and 4 doses of vaccine (29, 35, 36). Regarding T cell responses to VOCs, we observed that IMID patients retain robust T-cell mediated immunity against Omicron variants, consistent with studies demonstrating T cells are largely unaffected by Omicron variants and important for providing potent protection against emerging variants capable of escaping neutralization responses (18, 37–39). Future studies should investigate the humoral immunogenicity of the bivalent vaccines targeting BA.4 and BA.5 VOCs in IMID patients.

Analogous to humoral responses, IMID patients exhibited accelerated waning of T cell cytokine and cytotoxic molecule responses after a second dose of vaccine, which was corrected by a third dose. The decrease in responses by 3-4 months post-vaccination was no longer significant after three doses. IL-4 is of particular interest for vaccine-induced immunity as it is produced by T follicular helper cells (T_fh_) and promotes germinal center (GC) B cell proliferation, class-switching and differentiation (40). We noted that IL-4 levels increased with each of the first 3 doses of vaccine in healthy controls and IMID patients, with an additional increase in TNF IMID patients after the fourth dose. Whereas IL-4 levels correlated with the neutralization responses in non-TNF IMID patients after the second dose, this effect was only apparent in TNF IMID patients after the third dose, hinting at the mechanisms underlying the necessity of booster doses. As TNF inhibitors are known to disrupt the organization of germinal centers in rheumatoid arthritis patients (41), it is possible the cooperation between T_fh_ and GC B cells is disrupted in anti-TNF treated patients relative to other groups, hence a potential explanation for the lack of correlation of T cell IL-4 and Ab responses after dose 2. It is possible that by increasing the number of antigen-specific T and B cells and subsequently the cumulative magnitude of their functional responses, dose 3 may overcome this organizational deficit, allowing T_fh_ to provide help to B cells, leading to enhanced neutralizing Ab responses, although this remains speculative. A caveat of our study design is that we measured the total cytokine produced after T cell restimulation, but not the number of SARS-CoV-2-specific IL-4-producing T cells. Regardless, the results show that booster doses of vaccination are necessary to enhance the magnitude of T cell cytokine production.

This study has several limitations. Each study group (i.e., TNF IMID and non-TNF IMID) was heterogeneous in terms of disease type and specific treatment regimen, therefore we were underpowered to evaluate these effects within each group. Sample sizes declined for timepoints post third and fourth doses (T5-T8) of the vaccine because participants did not receive booster doses, withdrew from the study, or were excluded due to evidence of natural infection with SARS-CoV-2. Thus, after the fourth dose, it was difficult to draw conclusions about specific treatment groups. The latest timepoint assessed after each dose of vaccine was at the 3-4 months mark; we did not track long-term waning beyond that mark. Additionally, we did not assess hybrid immunity as we were limited by the low counts of vaccinated and infected patients. In Canada, fourth vaccine doses were offered to immunocompromised populations several months before they became available to healthy adults, resulting in a lack of healthy controls to compare to IMID patients for the post-dose four timepoints. Another limitation is the narrow dynamic range of the ELISA assays, meaning that saturating IgG levels below the limit of quantification could contribute to an underestimation of the effects of booster doses on the magnitude of responses and/or an underestimation of the deficits observed in TNF IMID patients compared to non-TNF IMID patients and healthy controls. Moreover, this study only assessed systemic immunity and did not examine mucosal immunity. Finally, the scope of the IMPACT study lies within investigating the immunogenicity of mRNA vaccines; our results do not make any inferences regarding vaccine efficacy in IMID patients.

Taken together, we present comprehensive data on the longitudinal course of vaccine-induced adaptive immunity in IMID patients, treated with systemic or targeted immune-modifying drugs. Our study suggests that repeated doses of vaccine prolong and broaden immune responses to SARS-CoV-2, supporting the recommendation for three and four-dose vaccine regimens in immunocompromised patients.

## Methods

### Study design and participants

In this prospective observational cohort study, we investigated the <u>IM</u>mune res<u>P</u>onse <u>A</u>fter <u>C</u>OVID-19 vaccination during maintenance <u>T</u>herapy in immune-mediated inflammatory diseases (IMPACT). The methods of the IMPACT study were previously reported in Refs (20, 21). In brief, we recruited adult participants, including healthy controls and patients diagnosed with one or more of the following IMIDs: inflammatory bowel disease, psoriasis, psoriatic arthritis, ankylosing spondylitis, rheumatoid arthritis, or hidradenitis suppurativa. IMID patients were untreated or treated with anti-IL-12/23 therapy, anti-IL-17 therapy, anti-IL-23 therapy, anti-tumor necrosis factor (TNF) therapy, methotrexate or azathioprine (MTX/AZA) monotherapy, or anti-TNF plus MTX/AZA combination therapy. Participants received one to four homologous or heterologous doses of BNT162b2 (Pfizer/BioNTech) or mRNA-1273 (Moderna) SARS-CoV-2 mRNA vaccines. Participants less than 18 years of age, those with prior SARS-CoV-2 infection, patients on oral steroids or B cell depletion agents, and those receiving non-mRNA SARS-CoV-2 vaccines were excluded.

### Sample collection

Blood samples were collected from each participant at up to 8 timepoints spanning pre and post one to four doses of vaccine, as defined in Table 1 -T1: pre-vaccination; T2: 2-4 weeks post dose 2; T3: 2-3 weeks post dose 2; T4: 3-4 months post dose 2; T5: 2-4 weeks post dose 3; T6: 3-4 months post dose 3; T7: 2-4 weeks post dose 4; T8: 3-4 months post dose 4. Plasma and peripheral blood mononuclear cells (PBMC) were isolated from blood (via centrifugation and using SepMate PBMC isolation tubes) for immunogenicity assessment.

### Immunogenicity assessment

Antibody (IgG) responses against coronavirus spike protein (S), receptor binding domain (RBD), and nucleocapsid (NP) were measured by automated ELISA (20). Data were compared to median convalescent values (anti-spike and anti-RBD IgG) for serum samples from 340 PCR-confirmed cases of COVID-19 cases 21-115 days after symptom onset (20). Spike-pseudotyped lentivirus neutralization assays were performed to assess neutralization capacity against WT SARS-CoV-2 and variants of concern (VOCs) including Omicron BA.1 and BA.5 (20). To assess SARS-CoV-2 specific T cell responses, PBMCs were stimulated for 48 hours with WT or Omicron BA.1 or BA.5 spike peptide pools (JPT Peptide Technologies). The release of 8 cytokines and cytotoxic molecules (IL-2, IL-4, IL-17A, IFNγ, Granzyme A, Granzyme B, sFasL, Perforin) in cell culture supernatants were measured using the LEGENDplex CD8/NK multiplex cytokine bead-based immunoassay (BioLegend) according to the manufacturer’s instructions. Results are reported after subtracting background values from negative control (DMSO stimulation) wells. More complete details are provided in Refs (20). *Statistics.* Data from timepoints T1-T5 were previously reported (Dayam *et al.* 2022, Cheung *et al.* 2022) and reanalyzed here (Figures 1-6) using additional statistical approaches. Least-squares linear regression models controlled for age, BMI, sex and vaccine type (Figure 1). Mixed-effects multivariate linear regression models controlled for age, BMI, sex and vaccine type (Figure 2-3, 5), including an interaction term between timepoint and study group. Differences in the effect of anti-TNF treatment by disease group were tested using linear regression models as described above, but with an interaction term between treatment with anti-TNF therapy and diagnosis with IBD (Figure 4). Associations between T cell responses (IL-2 and IL-4) in the 2-4 weeks following vaccination and neutralizing responses 3 months later were tested using regression models that controlled for age, BMI, sex, and vaccine type, and included an interaction term between the cytokine level and treatment with anti-TNF therapy (Figure 6). Samples provided by participants that presented evidence of a SARS-CoV-2 infection (seropositive for nucleocapsid IgG or self-reported a positive PCR or RAPID test) or were missing key covariates were excluded from analyses. A P value less than 0.05 was considered significant. Regression modeling was conducted in STATA/BE 17.0. Additional software utilized in creating graphics and/or statistical analyses includes the LEGENDplex Data Analysis Software Suite and R (version 4.2.1) with packages haven, ggpubr, and custom R scripts.

### Study approval

This study was approved by the ethics boards of the University of Toronto (REB protocol 27673), Mount Sinai Hospital/Sinai Health System (MSH REB 21-0022-E), University Health Network-Toronto Western Hospital division (REB 21-5096), and Women’s College Hospital (REB approval 2021-0023-E). Written informed consent was obtained from all participants prior to participation.

## Author contributions

MWC conducted T cell assays, analyzed data, conducted statistical analyses, prepared figures, drafted and edited the manuscript.

RMD conducted neutralization assays, analyzed antibody data, conducted statistical analysis, prepared figures and edited the manuscript.

JRS analyzed data, conducted statistical analysis, prepared figures, drafted and edited the manuscript.

MWC, RMD and JRS are co-first authors, authorship order was mutually agreed upon.

JCL designed and assisted with T cell assays.

GYCC provided overall project management including patient recruitment and validation of data records.

RLG contributed to study conception and design.

JMS and DP contributed to project management and patient recruitment.

DC, LA, SR, JDL, LJ, DG contributed to patient recruitment.

RL, VERC, MK assisted with PBMC preparation and/or T cell assays.

MD-B and GM handled samples and conducted ELISA assays.

VP and MSS contributed to study design, acquisition of funding, supervised clinical coordinators and patient recruitment, contributed to data interpretation and manuscript editing.

VC contributed to study design, acquisition of funding, supervision of patient recruitment, data analysis and edited the manuscript.

A-CG and THW contributed to study design, supervised the laboratory assays, analyzed data, acquired funding and wrote the manuscript.

## Funding

This project was supported by funding from the Public Health Agency of Canada, through the Vaccine Surveillance Reference group and the COVID-19 Immunity Task Force and by a donation from Juan and Stefania Speck. Additional funding was provided by Canadian Institutes of Health (CIHR)/COVID-Immunity Task Force (CITF) grants VR-1 172 711 and vs1-175545 (to THW and ACG), CIHR FDN-143250 (to THW) and FDN-143301 (to ACG), GA2-177716 (to VC, ACG and THW) and GA1-177703 (to ACG) and the CIHR rapid response network to SARS-CoV-2 variants, CoVaRR-Net (to ACG).

## Data Availability

All data produced in the present study are available upon reasonable request to the authors

## Acknowledgements

Thanks to Juan and Stefania Speck for their generous donations to the IMPACT study. We thank Birinder Ghumman for technical support and Natalie Simard for assistance in the flow cytometry facility. We thank Reuben Samson, Queenie Hu and W. Rod Hardy, and CoVaRR-Net for contributing the lentiviral particles for neutralization assays. We thank Dr. Karen Colwill, and Tulunay Tursun, Jenny Wang, Freda Qi and Adrian Pasculescu for help with generating and analyzing ELISA data. The equipment used for ELISA assays is housed in the Network Biology Collaborative Centre at the Lunenfeld-Tanenbaum Research Institute, a facility supported by Canada Foundation for Innovation funding (grant CFI 33474), by the Ontario Government, Genome Canada, and Ontario Genomics (OGI-139). Vinod Chandran is supported by a Pfizer Chair Research Award, Rheumatology, University of Toronto. David Croitoru is supported by the CSTP programme at the Department of Medicine, University of Toronto. Tania H. Watts holds the Canada ResearchChair in anti-viral immunity. Anne-Claude Gingras is the Canada Research Chair in Functional Proteomics and a pillar lead for CoVaRR-Net.

## Notes

### Author Declarations

Study approval. This study was approved by the ethics boards of the University of Toronto (REB protocol 27673), Mount Sinai Hospital/Sinai Health System (MSH REB 21-0022-E), University Health Network-Toronto Western Hospital division (REB 21-5096), and Women's College Hospital (REB approval 2021-0023-E). Written informed consent was obtained from all participants prior to participation.

