## Supplementary documents for "Third and fourth vaccine doses broaden and prolong immunity to SARS-CoV-2 in immunocompromised adult patients"

### Supplemental Figures

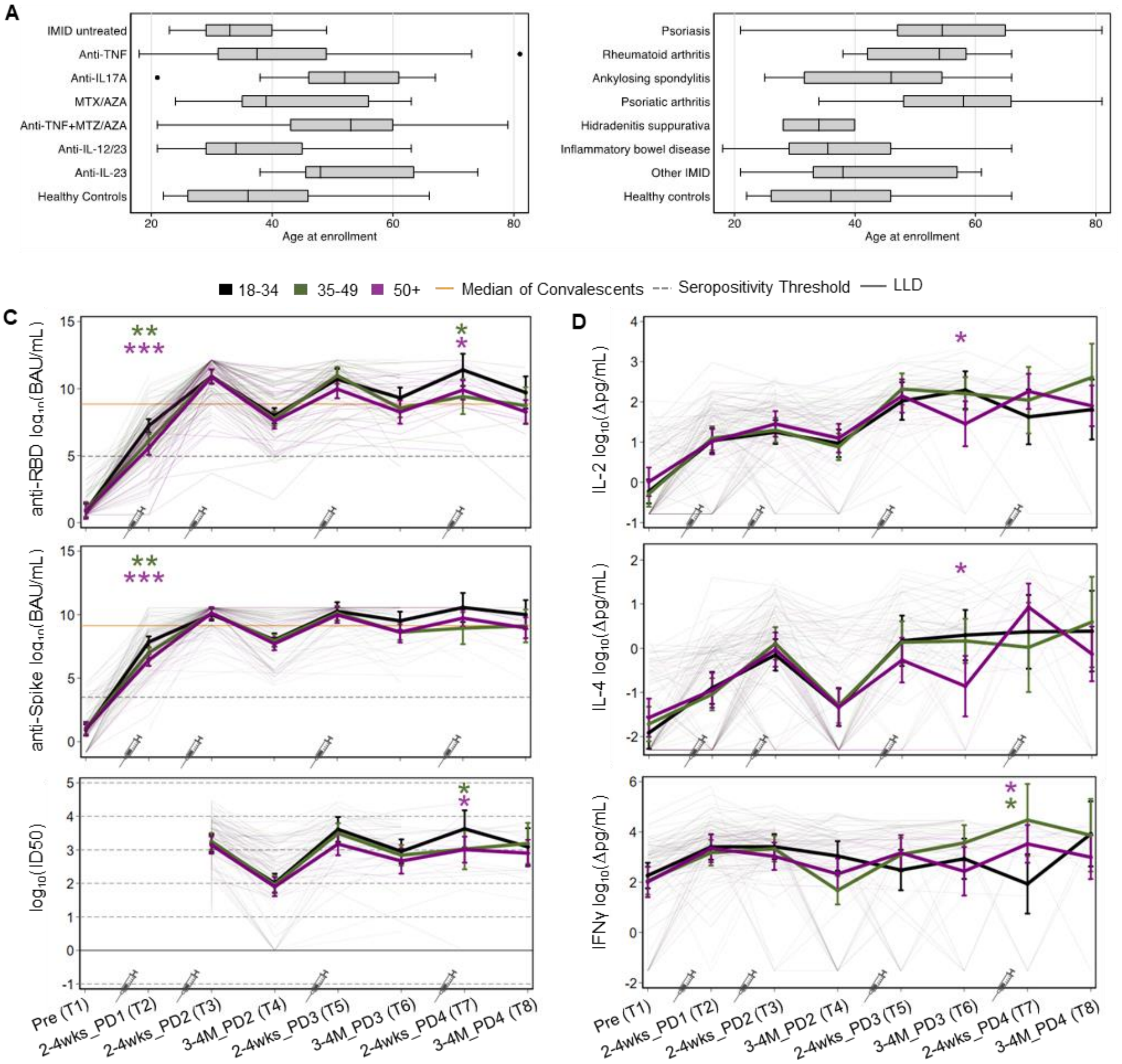


**Supplemental Figure 1. Effect of age on SARS-CoV-2 vaccine responses among IMID patients. (A)** The distribution of age at enrollment is shown by treatment group: N=9 IMID untreated, N=46 anti-TNF, N=12 anti-IL17A, N=9 MTX/AZA, N=19 anti-TNF+MTZ/AZA, N=29 anti-IL-12/23, N=12 anti-IL-23, and N=25 healthy controls. **(B)** Age distribution is also shown by diagnoses, which are not mutually exclusive: N= 38 psoriasis, N=8 rheumatoid arthritis, N=8 ankylosing spondylitis, N=31 psoriatic arthritis, N=2 hidradenitis suppurativa, N=86 inflammatory bowel disease, N=15 other IMID, N=25 healthy controls. **(C)** Longitudinal analyses of the effect of age on humoral responses across one to four doses of vaccine in IMID patients: anti-RBD and anti-spike IgG levels ($log_2$(BAU/mL)) and neutralization of Wildtype SARS-CoV-2 ($log_{10}$(ID50)). **(D)** Longitudinal analyses of the effect of age on cellular responses across one to four doses of vaccine in IMID patients: IL-2, IL-4, and IFNγ ($log_{10}$ (Δpg/mL)). **(C-D)** IMID patients are divided into three age groups: 18-34 (black), 35-49 (green), and 50+ (purple). Mixed-effects multivariate linear regression models controlled for BMI and sex with an interaction tern between timepoint and age group. An additional control for vaccine type was included in the regression models to compare responses between age groups: 18-34 vs 35-49 (green asterisks) and 18-34 vs 50+ (purple asterisks). *p<0.05; **p<0.01; ***p<0.001; Δ, background subtracted; pg/mL, picogram per milliliter; PD, post dose

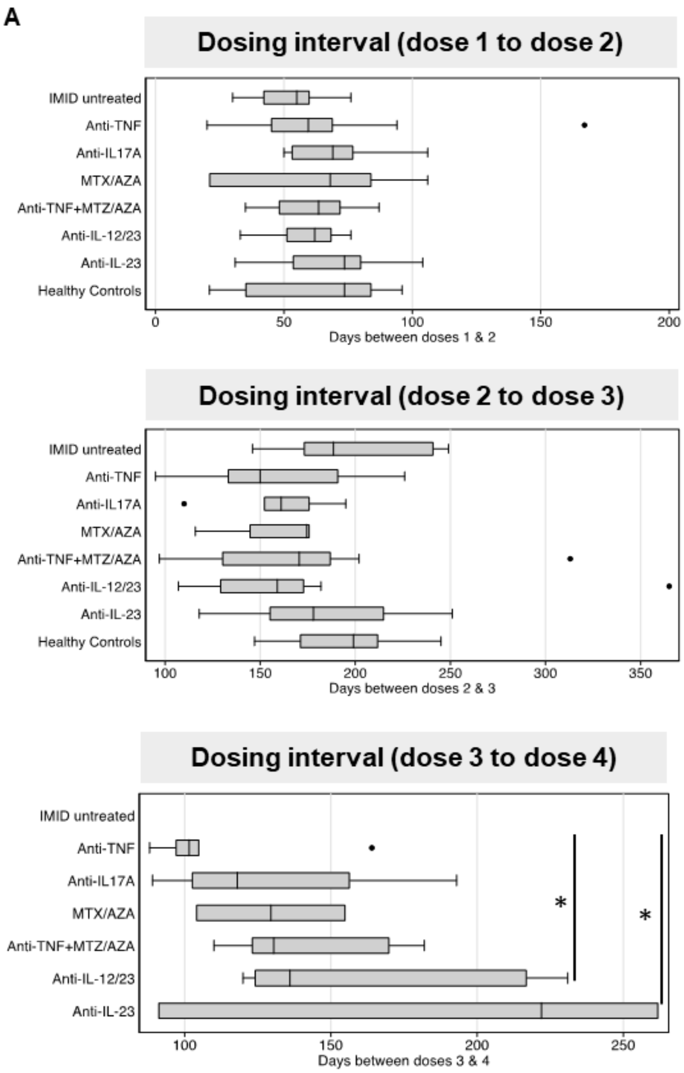


**Supplemental Figure 2. Dosing interval by treatment group.** Distribution of the days between dose1-dose2, dose2-dose3, and dose3-dose4 stratified by drug treatment group. *p<0.05

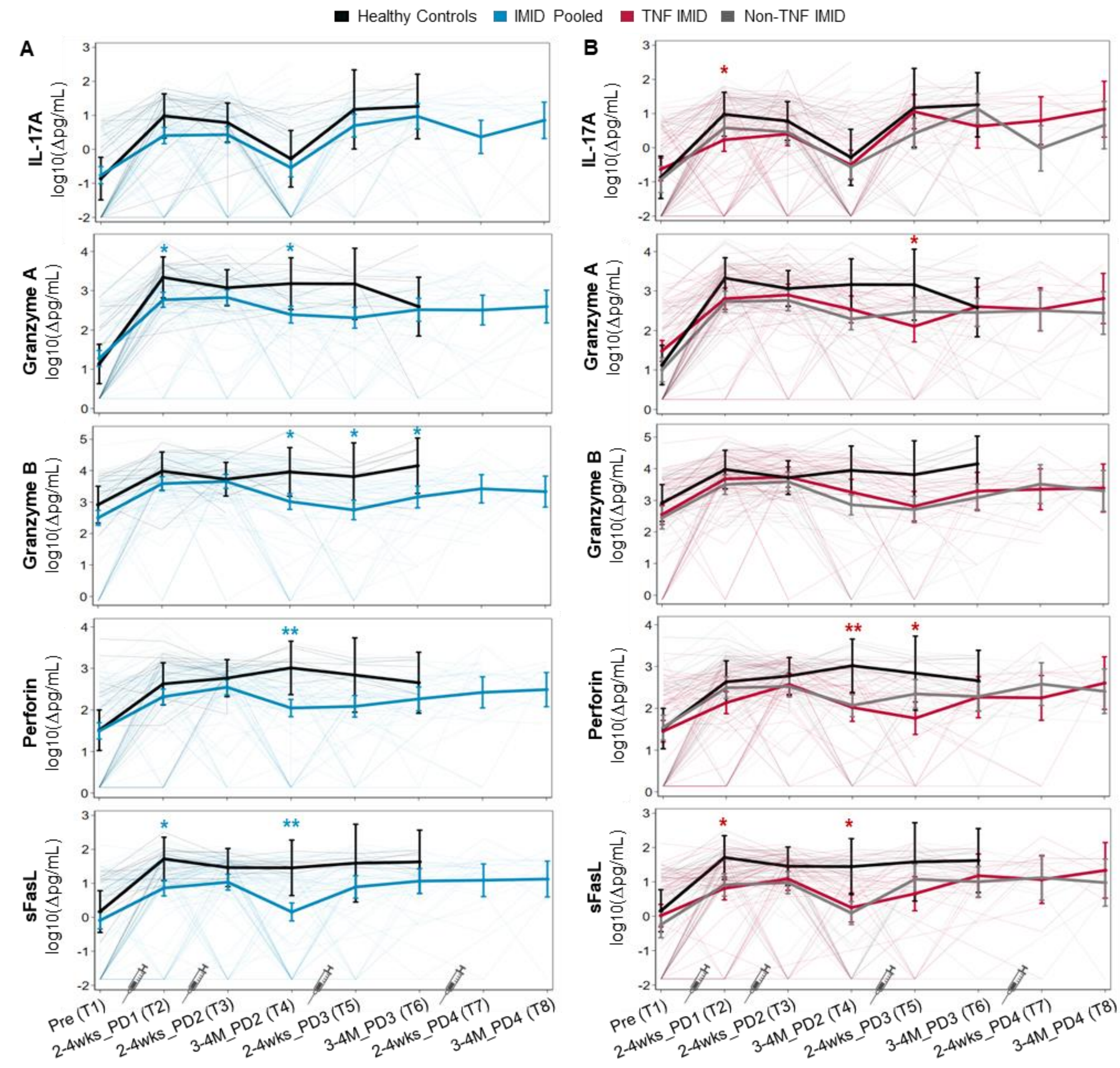


**Supplemental Figure 3. Longitudinal analyses of the magnitude of T cell cytokine and cytotoxic molecule responses pre and post one to four doses of SARS-CoV-2 mRNA vaccine.** Timepoints are defined in Table 1. **(A, B)** Predicted mean T cell cytokines and cytotoxic molecules IL-17A, Granzyme A, Granzyme B, sFasL, Perforin ($log_{10}$ (Δpg/mL)) from pre-vaccination to after four doses of vaccine. Mixed-effects linear regression models controlled for age, BMI, and sex (thick colored lines), with an interaction term between timepoint and group. Individual patients are plotted in thin lines. Groupings: healthy controls (black), IMID patients pooled (blue), TNF IMID (anti-TNF or anti-TNF+MTX/AZA treated) patients (red), and IMID patients not treated with anti-TNF (grey). Mixed-effects linear regression models with an additional control for vaccine type were used to compare groups: (A) IMID pooled vs healthy controls (blue asterisks); or (B) TNF IMID vs healthy controls (red asterisks). Sample sizes are listed in Supplemental Table 1. *p<0.05; **p<0.01; ***p<0.001; Δ, background subtracted; pg/mL, picogram per milliliter; PD, post dose

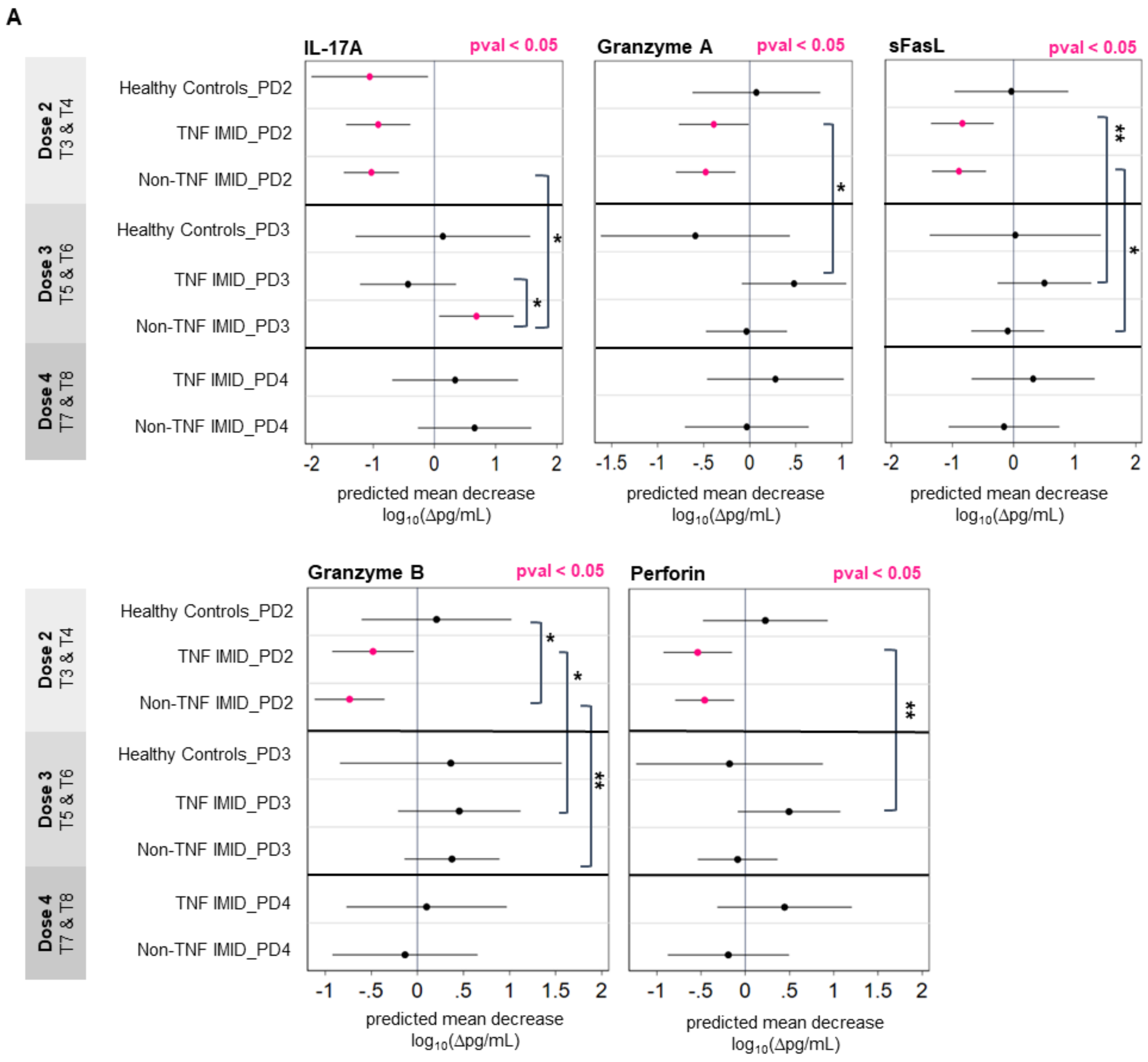


**Supplemental Figure 4. Longitudinal analyses of the stability of T cell cytokine and cytotoxic molecule responses post two to four doses of SARS-CoV-2 mRNA vaccine. (A)** Timepoints are defined in Table 1. Predicted mean decrease of cytokine and cytotoxic molecules IL-17A, Granzyme A, Granzyme B, sFasL, and Perforin ($log_{10}$ (Δpg/mL)) in healthy controls, TNF IMID, and IMID patients not treated with anti-TNF between 2-4 weeks and 3-4 months after dose 2 (decrease between T3 and T4), after dose 3 (decrease between T5 and T6), and after dose 4 (decrease between T7 and T8). Mixed-effects linear regression models controlled for age, BMI, sex, and vaccine type, with an interaction term between timepoint and group. Mean decrease values that are significant (pval<0.05) are colored in pink. Significant pairwise comparisons are indicated by asterisks and brackets. Sample sizes are listed in Supplemental Table 1. *p<0.05; **p<0.01; ***p<0.001; Δ: background subtracted; pg/mL: picogram per milliliter; PD: post dose

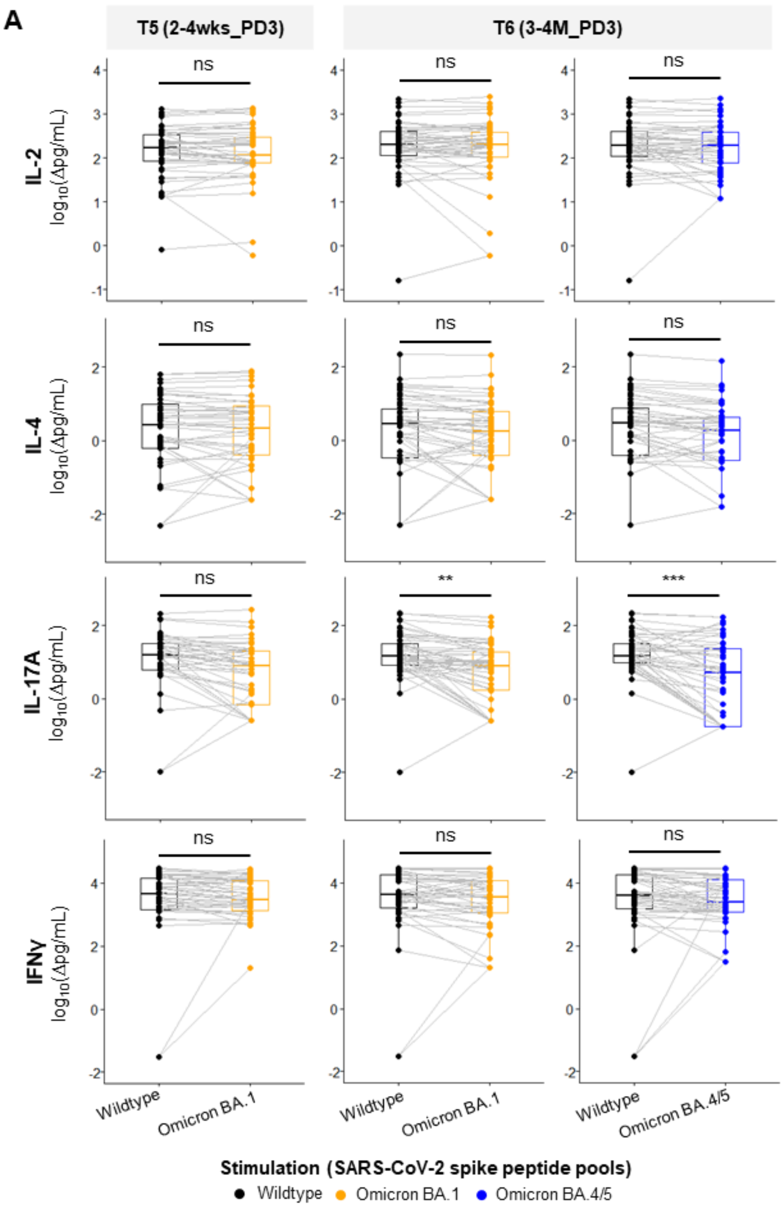


**Supplemental Figure 5. T cell cytokine responses to Wildtype SARS-Cov-2 and Omicron (BA.1, BA.4/5) after three doses of SARS-CoV-2 mRNA vaccine.** Timepoints are defined above each panel, as described in Table 1. **(A)** The release of cytokines in cell culture supernatant after 48 hours stimulation of PBMCs with SARS-CoV-2 spike peptide pools (lavender: Wildtype; orange: Omicron BA.1; blue: Omicron BA.4/5) were analyzed by LEGENDplex bead-based immunoassay. Depicted are responses in all participants (healthy and IMID) pooled, at two timepoints after the third dose of vaccine: T5 2-4wks_PD3 (2-4 weeks post dose 3) & T6 3-4M_PD3 (3-4 months post dose 3). Sample sizes are as follows: T5 (Wuhan vs Omicron BA.1), n = 46; T6 (Wuhan vs Omicron BA.1), n = 50; T6 (Wuhan vs Omicron BA.4/5), n = 52. Values are reported in pg/mL after subtracting background signal from wells containing PBMCs cultured with DMSO alone, as indicated by 'Δ'. Background subtracted values (Δpg/mL) were $log_{10}$ transformed for visualization purposes. Comparisons in paired IMID patients were conducted by paired Wilcoxon signed rank test after excluding healthy controls. *p<0.05, **p<0.01, ***p<0.001; ns, non-significant; PBMC, peripheral blood mononuclear cells; DMSO, dimethyl sulfoxide; pg/mL, picogram per millilitre; PD, post dose

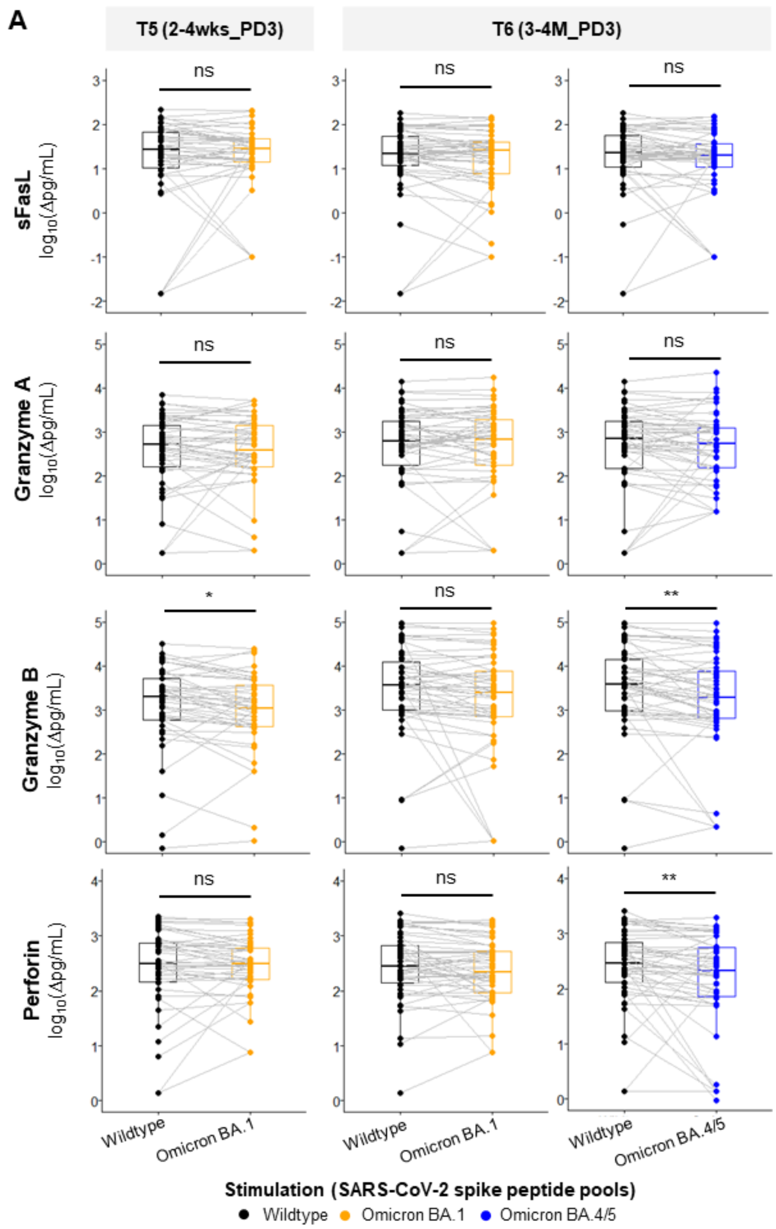


**Supplemental Figure 6. T cell cytotoxic responses to Wildtype SARS-CoV-2 and Omicron (BA.1, BA.4/5) after three doses of SARS-CoV-2 mRNA vaccine.** Timepoints are defined above each panel, as described in Table 1. **(A)** The release of cytotoxic molecules in cell culture supernatant after 48 hours stimulation of PBMCs with SARS-CoV-2 spike peptide pools (lavender: Wildtype; orange: Omicron BA.1; blue: Omicron BA.4/5) were analyzed by LEGENDplex bead-based immunoassay. Depicted are responses in all participants (healthy and IMID) pooled, at two timepoints after the third dose of vaccine: T5 2-4wks_PD3 (2-4 weeks post dose 3) & T6 3-4M_PD3 (3-4 months post dose 3). Sample sizes are as follows: T5 (Wuhan vs Omicron BA.1), n = 46; T6 (Wuhan vs Omicron BA.1), n = 50; T6 (Wuhan vs Omicron BA.4/5), n = 52. Values are reported in pg/mL after subtracting background signal from wells containing PBMCs cultured with DMSO alone, as indicated by 'Δ'. Background subtracted values (Δpg/mL) were $log_{10}$ transformed for visualization purposes. Comparisons in paired IMID patients were conducted by paired Wilcoxon signed rank test after excluding healthy controls. *p<0.05, **p<0.01, ***p<0.001; ns, non-significant; PBMC, peripheral blood mononuclear cells; DMSO, dimethyl sulfoxide; pg/mL, picogram per millilitre; PD, post dose

### Supplemental Tables

#### Supplemental Table S1. Samples sizes used in multivariate analyses

| | T1: Pre | T2: 2-4wks_PD1 | T3: 2-4wks_PD2 | T4: 3-4M_PD2 | T5: 2-4wks_PD3 | T6: 3-4M_PD3 | T7: 2-4wks_PD4 | T8: 3-4M_PD4 |
|---|---|---|---|---|---|---|---|---|
| **IgG** | | | | | | | | |
| **Healthy controls** | 15 | 17 | 21 | 11 | 5 | 6 | | |
| **IMID pooled** | **94** | **112** | **103** | **84** | **52** | **41** | **24** | **19** |
| **Non-TNF IMID** | **42** | **59** | **55** | **51** | **29** | **26** | **13** | **11** |
| Untreated | 9 | 9 | 9 | 9 | 5 | 6 | 0 | 0 |
| Anti-IL17A | 3 | 6 | 6 | 6 | 6 | 5 | 4 | 3 |
| MTX/AZA | 3 | 7 | 7 | 4 | 2 | 1 | 1 | 2 |
| Anti-IL12/23 | 26 | 27 | 23 | 22 | 11 | 11 | 6 | 5 |
| Anti-IL23 | 1 | 10 | 10 | 10 | 5 | 3 | 2 | 1 |
| **TNF IMID** | **52** | **53** | **48** | **33** | **23** | **15** | **11** | **8** |
| Anti-TNF | 37 | 37 | 34 | 23 | 16 | 9 | 5 | 3 |
| Anti-TNF + MTZ/AZA | 15 | 16 | 14 | 10 | 7 | 6 | 6 | 5 |
| **Neutralizing (Wildtype)** | | | | | | | | |
| **Healthy controls** | | | 21 | 9 | 5 | 6 | | |
| **IMID pooled** | | | **103** | **79** | **43** | **40** | **16** | **17** |
| **Non-TNF IMID** | | | **55** | **48** | **24** | **25** | **8** | **11** |
| Untreated | | | 9 | 9 | 5 | 5 | 0 | 0 |
| Anti-IL17A | | | 6 | 5 | 5 | 5 | 3 | 3 |
| MTX/AZA | | | 7 | 3 | 1 | 1 | 0 | 2 |
| Anti-IL12/23 | | | 23 | 22 | 8 | 11 | 4 | 5 |
| Anti-IL23 | | | 10 | 9 | 5 | 3 | 1 | 1 |
| **TNF IMID** | | | **48** | **31** | **19** | **15** | **8** | **6** |
| Anti-TNF | | | 34 | 22 | 14 | 9 | 4 | 2 |
| Anti-TNF + MTZ/AZA | | | 14 | 9 | 5 | 6 | 4 | 4 |
| **T cell cytokines and cytotoxic molecules** | | | | | | | | |
| **Healthy controls** | 14 | 13 | 17 | 8 | 4 | 6 | | |
| **IMID pooled** | **85** | **95** | **96** | **75** | **49** | **39** | **25** | **20** |
| **Non-TNF IMID** | **36** | **48** | **50** | **46** | **27** | **26** | **14** | **11** |
| Untreated | 9 | 9 | 9 | 9 | 5 | 6 | 0 | 0 |

| | | | | | | | | |
|---|---|---|---|---|---|---|---|---|
| Anti-IL17A | 3 | 5 | 4 | 5 | 6 | 5 | 4 | 3 |
| MTX/AZA | 3 | 5 | 6 | 3 | 1 | 1 | 1 | 2 |
| Anti-IL12/23 | 20 | 20 | 22 | 21 | 10 | 11 | 7 | 5 |
| Anti-IL23 | 1 | 9 | 9 | 8 | 5 | 3 | 2 | 1 |
| **TNF IMID** | **49** | **47** | **46** | **29** | **22** | **13** | **11** | **9** |
| Anti-TNF | 36 | 33 | 32 | 20 | 14 | 7 | 5 | 4 |
| Anti-TNF + MTZ/AZA | 13 | 14 | 14 | 9 | 8 | 6 | 6 | 5 |

Bolded values represent sums of the rows below (i.e., IMID pooled totals are the sum of Non-TNF IMID and TNF IMID totals).
Abbreviations: AZA: azathioprine; IL: Interleukin; IMID: Immune-Mediated Inflammatory Diseases; M: months; MTX: Methotrexate; PD: Post dose; TNF: Tumor necrosis factor; wks: weeks

**Supplemental Table S2. Sample sizes used in comparisons of neutralization of Wildtype SARS-CoV-2 versus Omicron BA.1/BA.5.**

| Neutralizing | T4: 3-4M_PD2 | T5: 2-4wks_PD3 | T6: 3-4M_PD3 | T7: 2-4wks_PD4 | T8: 3-4M_PD4 |
|---|---|---|---|---|---|
| **Wildtype vs Omicron BA.1** | | | | | |
| Healthy controls | 9 | 5 | 6 | | |
| Non-TNF IMID | 48 | 24 | 25 | 8 | 11 |
| TNF IMID | 32 | 19 | 15 | 9 | 6 |
| **Wildtype vs Omicron BA.5** | | | | | |
| Healthy controls | | 5 | 6 | | |
| Non-TNF IMID | | 24 | 25 | 8 | 11 |
| TNF IMID | 4 | 19 | 15 | 9 | 6 |

**Supplemental Table S3. Anti-spike and anti-RBD IgG levels**

| | Anti-spike - $log_2$(BAU/mL) | | | Anti-RBD - $log_2$(BAU/mL) | | |
|---|---|---|---|---|---|---|
| | **Healthy controls** | **Non-TNF IMID** | **Anti-TNF** | **Healthy controls** | **Non-TNF IMID** | **Anti-TNF** |
| **T1: Pre - N** | 15 | 42 | 52 | 15 | 42 | 52 |
| Median (IQR) | 0.8 (-0.8 - 1.5) | 0.7 (0.3 - 1.3) | 0.8 (0.4 - 1.6) | 0.6 (0.6 - 0.6) | 0.6 (0.6 - 0.6) | 0.6 (0.6 - 0.6) |
| Upper limit - N (%) | 0 (0.0) | 0 (0.0) | 0 (0.0) | 0 (0.0) | 0 (0.0) | 0 (0.0) |
| **T2: 2-4wks_PD1 - N** | 17 | 59 | 53 | 16 | 58 | 53 |
| Median (IQR) | 8.3 (7.9 - 8.9) | 7.6 (6.5 - 8.8) | 6.4 (5.4 - 7.2) | 7.6 (7.0 - 8.3) | 7.2 (6.4 - 7.8) | 6.1 (4.8 - 7.1) |
| Upper limit - N (%) | 0 (0.0) | 2 (3.4) | 0 (0.0) | 0 (0.0) | 0 (0.0) | 0 (0.0) |
| **T3: 2-4wks_PD2 - N** | 21 | 55 | 48 | 21 | 55 | 48 |
| Median (IQR) | 10.6 (10.6 - 10.6) | 10.6 (10.6 - 10.6) | 10.5 (9.2 - 10.6) | 11.7 (10.6 - 12.1) | 12.0 (11.3 - 12.1) | 10.5 (9.1 - 11.9) |
| Upper limit - N (%) | 17 (81.0) | 45 (81.8) | 24 (50.0) | 9 (42.9) | 25 (45.5) | 10 (20.8) |
| **T4: 3-4M_PD2 - N** | 11 | 51 | 33 | 11 | 51 | 33 |
| Median (IQR) | 9.7 (9.1 - 10.0) | 9.2 (7.9 - 9.7) | 5.7 (5.4 - 7.3) | 9.5 (9.2 - 10.5) | 8.7 (7.5 - 10.1) | 6.7 (5.1 - 7.2) |
| Upper limit - N (%) | 0 (0.0) | 2 (3.9) | 0 (0.0) | 0 (0.0) | 0 (0.0) | 0 (0.0) |
| **T5: 2-4wks_PD3 - N** | 5 | 29 | 23 | 5 | 29 | 23 |
| Median (IQR) | 10.6 (10.6 - 10.6) | 10.6 (10.5 - 10.6) | 10.6 (9.2 - 10.6) | 11.6 (11.3 - 11.7) | 11.2 (10.4 - 11.8) | 10.5 (8.4 - 11.5) |
| Upper limit - N (%) | 5 (100.0) | 21 (72.4) | 13 (56.5) | 1 (20.0) | 4 (13.8) | 2 (8.7) |
| **T6: 3-4M_PD3 - N** | 6 | 26 | 15 | 6 | 26 | 14 |
| Median (IQR) | 10.5 (9.9 - 10.6) | 9.6 (9.0 - 10.5) | 8.1 (6.1 - 9.2) | 10.0 (9.8 - 10.8) | 9.3 (8.5 - 10.3) | 8.1 (7.2 - 8.6) |
| Upper limit - N (%) | 3 (50.0) | 6 (23.1) | 1 (6.7) | 0 (0.0) | 1 (3.8) | 0 (0.0) |
| **T7: 2-4wks_PD4 - N** | | 13 | 11 | | 13 | 11 |
| Median (IQR) | | 10.6 (10.6 - 10.6) | 9.1 (8.2 - 10.6) | | 11.0 (10.7 - 11.7) | 9.0 (8.6 - 10.4) |
| Upper limit - N (%) | | 11 (84.6) | 3 (27.3) | | 2 (15.4) | 1 (9.1) |
| **T8: 3-4M_PD4 - N** | | 11 | 8 | | 11 | 8 |
| Median (IQR) | | 9.8 (9.4 - 10.6) | 8.6 (6.7 - 9.6) | | 9.1 (8.5 - 10.7) | 7.7 (7.1 - 9.5) |
| Upper limit - N (%) | | 4 (36.4) | 1 (12.5) | | 0 (0.0) | 0 (0.0) |

Abbreviations: BAU/mL: Binding antibody units per milliliter; IMID: Immune-Mediated Inflammatory Diseases; IQR: Inter-quartile range; M: months; PD: Post dose; TNF: Tumor necrosis factor; wks: weeks

**Supplemental Table S4. Neutralization titers**

| | Wild Type Neutralization - $Log_{10}$(ID50) | | | Omicron BA.1 Neutralization - $Log_{10}$(ID50) | | | Omicron BA.5 Neutralization - $Log_{10}$(ID50) | | |
|---|---|---|---|---|---|---|---|---|---|
| | **Healthy controls** | **Non-TNF IMID** | **Anti-TNF** | **Healthy controls** | **Non-TNF IMID** | **Anti-TNF** | **Healthy controls** | **Non-TNF IMID** | **Anti-TNF** |
| **T3: 2-4wks_PD2 - N** | 21 | 55 | 48 | | | | | | |
| Median (IQR) | 3.6 (3.2 - 3.9) | 3.5 (3.3 - 3.9) | 2.9 (2.4 - 3.4) | | | | | | |
| Lower limit - N (%) | 0 (0.0) | 0 (0.0) | 1 (2.1) | | | | | | |
| **T4: 3-4M_PD2 - N** | 9 | 48 | 31 | 9 | 48 | 31 | | | |
| Median (IQR) | 3.2 (3.0 - 3.2) | 2.5 (2.2 - 2.8) | 1.6 (0.0 - 2.0) | 2.1 (1.3 - 2.2) | 1.1 (0.0 - 1.7) | 0.0 (0.0 - 0.0) | | | |
| Lower limit - N (%) | 0 (0.0) | 1 (2.1) | 8 (25.8) | 1 (11.1) | 17 (35.4) | 25 (80.6) | | | |
| **T5: 2-4wks_PD3 - N** | 5 | 24 | 19 | 5 | 24 | 19 | 5 | 24 | 19 |
| Median (IQR) | 3.8 (3.6 - 3.9) | 3.5 (3.3 - 3.6) | 3.6 (2.9 - 3.9) | 2.9 (2.8 - 3.3) | 2.5 (2.3 - 2.7) | 2.5 (0.9 - 3.0) | 2.9 (2.6 - 3.1) | 2.4 (2.2 - 2.6) | 2.2 (1.8 - 2.7) |
| Lower limit - N (%) | 0 (0.0) | 0 (0.0) | 0 (0.0) | 0 (0.0) | 0 (0.0) | 4 (21.1) | 0 (0.0) | 0 (0.0) | 1 (5.3) |
| **T6: 3-4M_PD3 - N** | 6 | 25 | 15 | 6 | 25 | 15 | 6 | 25 | 15 |
| Median (IQR) | 3.7 (3.4 - 3.9) | 3.1 (2.9 - 3.2) | 2.7 (2.2 - 3.1) | 2.6 (2.5 - 3.1) | 2.0 (1.8 - 2.3) | 1.6 (0.0 - 1.8) | 2.3 (2.2 - 2.6) | 1.8 (1.4 - 2.1) | 0.0 (0.0 - 1.6) |
| Lower limit - N (%) | 0 (0.0) | 0 (0.0) | 0 (0.0) | 0 (0.0) | 0 (0.0) | 5 (33.3) | 0 (0.0) | 3 (12.0) | 8 (53.3) |
| **T7: 2-4wks_PD4 - N** | | 8 | 8 | | 8 | 8 | | 8 | 8 |
| Median (IQR) | | 3.6 (3.1 - 3.8) | 3.0 (2.2 - 3.5) | | 2.5 (1.3 - 2.8) | 1.2 (1.0 - 2.3) | | 2.5 (2.0 - 3.0) | 1.6 (1.0 - 2.3) |
| Lower limit - N (%) | | 0 (0.0) | 1 (12.5) | | 1 (12.5) | 1 (12.5) | | 0 (0.0) | 1 (12.5) |
| **T8: 3-4M_PD4 - N** | | 11 | 6 | | 11 | 6 | | 11 | 6 |
| Median (IQR) | | 3.4 (2.9 - 3.6) | 2.3 (2.1 - 2.9) | | 2.2 (1.0 - 2.4) | 1.3 (1.2 - 1.6) | | 2.1 (1.6 - 2.7) | 1.2 (0.0 - 1.6) |
| Lower limit - N (%) | | 0 (0.0) | 0 (0.0) | | 1 (9.1) | 1 (16.7) | | 1 (9.1) | 2 (33.3) |

Abbreviations: ID50: 50% inhibitory dose; IMID: Immune-Mediated Inflammatory Diseases; IQR: Inter-quartile range; M: months; PD: Post dose; TNF: Tumor necrosis factor; wks: weeks

**Supplemental Table S5. Comparisons (p-value) of peak humoral and cellular responses after successive doses of vaccination**

| **Humoral Responses** | | | | |
|---|---|---|---|---|
| **Anti-spike IgG - $log_2$(BAU/mL)** | **Healthy Controls** | **IMID Pooled** | **TNF IMID** | **Non-TNF IMID** |
| Dose 1 vs Dose 2 (T2 vs T3) | 0.000 | 0.000 | 0.000 | 0.000 |
| Dose 2 vs Dose 3 (T3 vs T5) | 0.563 | 0.855 | 0.740 | 0.487 |
| Dose 3 vs Dose 4 (T5 vs T7) | | 0.488 | 0.481 | 0.688 |
| **Anti-RBD IgG - $log_2$(BAU/mL)** | | | | |
| Dose 1 vs Dose 2 (T2 vs T3) | 0.000 | 0.000 | 0.000 | 0.000 |
| Dose 2 vs Dose 3 (T3 vs T5) | 0.822 | 0.105 | 0.366 | 0.187 |
| Dose 3 vs Dose 4 (T5 vs T7) | | 0.312 | 0.283 | 0.702 |
| **Neutralization of Wildtype - $log_{10}$(ID50)** | | | | |
| Dose 1 vs Dose 2 (T2 vs T3) | | | | |
| Dose 2 vs Dose 3 (T3 vs T5) | | 0.019 | 0.005 | 0.353 |
| Dose 3 vs Dose 4 (T5 vs T7) | | 0.111 | 0.071 | 0.484 |
| **Cellular Responses** | | | | |
| **IL-2 - $log_{10}$(Δpg/mL)** | **Healthy Controls** | **IMID Pooled** | **TNF IMID** | **Non-TNF IMID** |
| Dose 1 vs Dose 2 (T2 vs T3) | 0.081 | 0.026 | 0.003 | 0.846 |
| Dose 2 vs Dose 3 (T3 vs T5) | 0.195 | 0.000 | 0.000 | 0.000 |
| Dose 3 vs Dose 4 (T5 vs T7) | | 0.583 | 0.694 | 0.237 |
| **IL-4 - $log_{10}$(Δpg/mL)** | | | | |
| Dose 1 vs Dose 2 (T2 vs T3) | 0.013 | 0.000 | 0.000 | 0.000 |
| Dose 2 vs Dose 3 (T3 vs T5) | 0.141 | 0.861 | 0.554 | 0.775 |
| Dose 3 vs Dose 4 (T5 vs T7) | | 0.018 | 0.027 | 0.268 |
| **IFNγ - $log_{10}$(Δpg/mL)** | | | | |
| Dose 1 vs Dose 2 (T2 vs T3) | 0.449 | 0.395 | 0.365 | 0.748 |
| Dose 2 vs Dose 3 (T3 vs T5) | 0.609 | 0.233 | 0.110 | 0.838 |
| Dose 3 vs Dose 4 (T5 vs T7) | | 0.776 | 0.763 | 0.852 |

Abbreviations: ID50: 50% inhibitory dose; BAU/ml: binding antibody units per milliliter; Δ: delta, background subtracted; pg/mL: picogram per milliliter; colored in red: p-value < 0.05
